# Diagnostic performance of non-invasive fibrosis markers for chronic hepatitis B in sub-Saharan Africa: a Bayesian individual patient data meta-analysis

**DOI:** 10.1101/2022.03.18.22272415

**Authors:** Asgeir Johannessen, Alexander J. Stockdale, Marc Y.R. Henrion, Edith Okeke, Moussa Seydi, Gilles Wandeler, Mark Sonderup, C. Wendy Spearman, Michael Vinikoor, Edford Sinkala, Hailemichael Desalegn, Fatou Fall, Nicholas Riches, Pantong Davwar, Mary Duguru, Tongai Maponga, Jantjie Taljaard, Philippa C. Matthews, Monique Andersson, Roger Sombie, Yusuke Shimakawa, Maud Lemoine

**Affiliations:** Department of Infectious Diseases, Vestfold Hospital, Tønsberg, Norway; Institute of Clinical Medicine, University of Oslo, Oslo, Norway; Department of Clinical Infection, Microbiology and Immunology, Institute of Infection, Veterinary and Ecological Sciences, University of Liverpool, Liverpool, United Kingdom; Malawi-Liverpool-Wellcome Trust Clinical Research Programme, Blantyre, Malawi; Department of Clinical Sciences, Liverpool School of Tropical Medicine, Liverpool,UK; Faculty of Medical Sciences, University of Jos, Jos, Nigeria; Service de Maladies Infectieuses et Tropicales, Centre Regional de Recherche et de Formation, Centre Hospitalier National Universitaire de Fann, Dakar, Senegal; Institute of Social and Preventive Medicine, University of Bern, Switzerland; Division of Hepatology, Department of Medicine, Faculty of Health Sciences, University of Cape Town, Cape Town, South Africa; Department of Internal Medicine, University of Zambia, Lusaka, Zambia; University of Alabama at Birmingham, Birmingham, Alabama, USA; Medical Department, St. Paul’s Hospital Millennium Medical College, Addis Ababa, Ethiopia; Department of Hepatology and Gastroenterology, Hopital Principal de Dakar, Dakar, Senegal; Division of Medical Virology, Stellenbosch University Faculty of Medicine and Health Sciences, Cape Town, South Africa; Division of Infectious Diseases, Department of Medicine, Tygerberg Hospital and Stellenbosch University, Cape Town, South Africa; Nuffield Department of Medicine, University of Oxford, Oxford, UK; The Francis Crick Institute, London, United Kingdom; University College London, London, United Kingdom; Yalgado Ouédraogo University Hospital Center, Ouagadougou, Burkina Faso; Unité d’Epidémiologie des Maladies Emergentes, Institut Pasteur, Paris, France; Department of Metabolism, Digestion and Reproduction, Imperial College London, London, UK

## Abstract

**Objective:** In sub-Saharan Africa, hepatitis B is the principal cause of liver disease. Non-invasive biomarkers of liver fibrosis are needed to identify patients requiring antiviral treatment. We assessed aspartate aminotransferase-to-platelet ratio index (APRI), gamma-glutamyl transferase-to-platelet ratio (GPR) and FIB-4 to diagnose significant fibrosis and cirrhosis in an individual patient data (IPD) meta-analysis.

**Design:** In total, 3,549 patients from 12 cohorts of HBsAg positive individuals in 8 sub-Saharan African countries were included. Transient elastography was used as a reference test for cirrhosis (>12.2 kPa), excluding patients who were pregnant, had hepatitis C, D, or HIV co-infection, were on hepatitis B therapy, or had acute hepatitis. A bivariate Bayesian IPD model was fitted with patient-level covariates and study-level random effects.

**Results:** APRI and GPR had the best discriminant performance (area under receiver operating curve 0.81 and 0.82) relative to FIB-4 (0.77) for cirrhosis. The World Health Organization (WHO) recommended APRI threshold of ≥2.0 was associated with a sensitivity and specificity (95% credible interval) of 16.5% (12.5-20.5) and 99.5% (99.2-99.7) for cirrhosis. For APRI, we identified an optimised rule-in threshold for cirrhosis (cut-off 0.65) with a sensitivity and specificity of 56.2% (50.5-62.2) and 90.0% (89.0-91.0), and an optimised rule-out threshold (cut-off 0.36) with a sensitivity and specificity of 80.6% (76.1-85.1) and 64.3% (62.8-65.8).

**Conclusions:** The WHO recommended APRI threshold of 2.0 is too high to diagnose cirrhosis in sub-Saharan Africa. We identified new and optimised rule-in and rule-out thresholds for cirrhosis, with direct consequences for treatment guidelines in this setting.

## INTRODUCTION

Worldwide, an estimated 296 million people are living with chronic hepatitis B (CHB).^1^ The natural course of infection is variable, ranging from an inactive carrier state with an excellent long-term prognosis, to progressive hepatic necroinflammation leading to cirrhosis and/or hepatocellular carcinoma (HCC).^2^ Antiviral therapy effectively reduces the risk of these complications^3,4^, and the challenge in clinical practice is to identify patients at risk of progressive liver disease and to start timely antiviral therapy.

International treatment guidelines recommend antiviral therapy for patients with cirrhosis, or with elevated hepatitis B virus (HBV) viral load and significant liver fibrosis or inflammation.^5-8^ Cirrhosis can be diagnosed clinically only in an advanced (i.e. decompensated) phase. Earlier stages of liver fibrosis are traditionally assessed by liver biopsy, which over the past decade has been largely replaced by transient elastography (TE). In resource-limited settings, however, these fibrosis assessment tools are rarely available, and management of CHB patients relies on clinical assessment alone.

The World Health Organization’s (WHO) first guidelines for CHB published in 2015 recommended the use of non-invasive fibrosis markers based on low-cost routinely available laboratory tests in resource-limited settings, and this was adapted into national guidelines in many low- and middle-income countries. Specifically, the aspartate aminotransferase-to-platelet ratio index (APRI) at a threshold of 2.0 was recommended to identify patients with cirrhosis in the WHO 2015 guidelines, although the lack of data from sub-Saharan Africa (sSA) was acknowledged as an important knowledge gap.^8^ APRI, FIB-4 and most other fibrosis markers were developed and validated in Caucasian and Asian cohorts,^9,10^ where HBV genotypes, environmental exposures, endemic infections, and human and viral genetic factors differ from sSA. Indeed, emerging data from sSA have shown that the WHO recommended APRI threshold may have poor sensitivity in this setting for the diagnosis of cirrhosis.^11-13^

With a population size of more than one billion, and with 82 million people living with CHB, sSA is in desperate need of locally-adapted treatment guidelines for CHB that consider operational constraints and potential differences in the natural history of infection in the region.^14^ Guidelines need to consider the high prevalence of CHB and the significant resource implications to effectively monitor and make treatment decisions for patients in over-stretched health systems. To meet this demand, and to fill the knowledge gaps identified by the first WHO guidelines, we established HEPSANET, a collaborative network of 12 hepatitis B cohorts in sSA. In the present analysis we present an individual patient data meta-analysis of the diagnostic performance of APRI, FIB-4 and another low-cost biomarker, the gamma-glutamyl transferase-to-platelet ratio (GPR)^13^, among patients in the HEPSANET collaboration using TE as a reference test. TE is a widely-used clinical tool in high-income countries and has prognostic properties for predicting liver-related events and death.^15^ Numerous cross-sectional studies have compared it with liver biopsy, which has historically been considered a “gold standard” but is not feasible to perform in routine care in sSA due to infrastructure and resource limitations. Meta-analysis has shown a favourable correlation between TE-based classification and liver biopsy, with an area under the receiver operating curve (AUROC) of 0.88 for F≥2 and 0.93 for F4, respectively.^16^ Liver biopsy is also associated with limitations due to spectrum bias and sampling error, and it has been shown that AUROC >0.90 is unachievable even for a perfect surrogate marker.^17^ By pooling data from centres across sSA we believe findings from this analysis can be generalized to settings throughout the region.

## MATERIAL AND METHODS

### Systematic review for relevant cohorts

We searched PubMed, Scopus, the African Index Medicus and African Journals Online for articles evaluating individuals with chronic HBV infection in sSA published until on 6^th^ October 2020. Search terms included synonyms of hepatitis B, transient elastography or liver biopsy and the countries of sub-Saharan Africa (Appendix 1). No language restrictions were applied. Two investigators independently screened all identified articles and reviewed potentially eligible full-text articles for eligibility.

We included studies reporting on hepatitis B surface antigen (HBsAg) positive individuals aged ≥13 years living in sub-Saharan Africa who had pre-therapy evaluation with fasting (>2 hours) transient elastography and measurement of alanine aminotransferase (ALT), aspartate aminotransferase (AST) and platelet count. TE and blood tests were ideally performed on the same day but an interval of up to 3 months was accepted. We excluded studies reporting on patients originating in sSA who migrated outside Africa. Two investigators (AS and NR) independently extracted the variables listed in Appendix 2 and evaluated the risk of bias among included studies using the QUADAS-2 assessment tool with disagreement resolved by consensus.^18^

### Individual patient data

We contacted authors of all publications which met our study inclusion criteria, as well as researchers active in this area, to share individual patient data. For the current analysis, we excluded patients who had received antiviral treatment for HBV within the preceding six months, or who had started therapy for at least 7 days prior to the time of evaluation, patients with co-infection with hepatitis C (anti-HCV) or hepatitis D (anti-HDV) or human immunodeficiency virus (HIV), pregnant women, patients with ALT or AST exceeding 5 times the upper limit of normal (in accordance with European Association for the Study of the Liver reliability recommendations),^19^ and those with suspected or confirmed hepatocellular carcinoma. Centre-specific definitions were used to categorise patients with hazardous alcohol consumption as described in appendix 3; the majority used the WHO AUDIT tool.^20^

### Primary outcome and reference test

We assessed the diagnostic performance characteristics of non-invasive tests for the diagnosis of significant liver fibrosis and cirrhosis in CHB patients in sSA. Fasting TE was used as the reference test with cut-offs of 7.9 kPa and 12.2 kPa used to define significant fibrosis (equivalent to METAVIR ≥F2) and cirrhosis (F4) respectively, according to a meta-analysis of 4,386 CHB patients with cross-sectional comparisons with liver biopsies.^16^ Sensitivity analyses were used to assess an alternative threshold for cirrhosis of 9.5 kPa derived from a study of 135 CHB patients from The Gambia who underwent both TE and liver biopsy.^13^ In this study, the total number of cirrhotic patients was only 20 so the latter cut-off must be interpreted with caution.

We assessed the following biomarkers:

i. APRI = [(AST (U/L) / upper limit of normal (ULN) of AST range) x 100] / platelet count (×10^9^/L);^9^
ii. GPR = GGT (U/L) / ULN of GGT range / platelet count (×10^9^/L);^13^
iii. FIB-4 = (age (years) × AST (U/L)) / (platelet count (×10^9^/L) x (ALT (U/L))^1/2^);^10^
iv. ALT (U/L), as a standalone marker.

An ULN of 40 U/L for AST and 61 U/L for GGT were used in the models.

First, we evaluated the WHO recommended rule-in thresholds for APRI (at 1.5 for ≥F2 and 2.0 for F4 respectively).^8^ Proposed thresholds for GPR and FIB-4 derived from The Gambia were evaluated.^13^ We then developed rule-in thresholds, choosing cut-offs with specificity of at least 90%, and rule-out thresholds defined by a sensitivity of at least 80%. Finally, we assessed thresholds defined by Youden’s J, which maximises the sum of sensitivity and specificity.

### Statistical analysis

To calculate sensitivity and specificity, data were pooled using a single-stage individual patient data (IPD) meta-analysis approach. We used a bivariate Bayesian random-effects meta-analysis model for sensitivity and specificity using patient-level covariates with study-level random effects to account for anticipated variability between sites.^21^ Details of the model are provided in Appendix 4.

### Validation

We validated the model using bootstrap resampling. We obtained 500 bootstrap samples from the original dataset, then fitted the model and estimated rule-in, rule-out and Youden thresholds, sensitivity and specificity at different thresholds, and overall AUROC. Validation results for the APRI model for cirrhosis are shown in Appendix 5. Model parameters show unimodal distributions with, in general, narrow spread indicating good stability of parameter estimates. The figure for the APRI rule-in (specificity ≥ 90%) threshold appears to show a bimodal distribution. However, this is an artefact of the grid search we performed for optimal thresholds: 0.65 and 0.74 are consecutive values, so the validation shows that the bootstrapped estimates only switch between 2 consecutive values of the grid search.

### Sensitivity analyses

We evaluated the model with several sensitivity analyses. We first assessed the effect of excluding patients with ascites diagnosed clinically or radiologically, for whom the probability of cirrhosis is high and who would be likely to receive HBV treatment, regardless of biomarker findings. Second, we assessed the effect of using sex-specific upper limits of normal for AST in calculating APRI, as proposed in the WHO 2015 guidelines (male 30 U/L; female 19 U/L).^8^ Third, we assessed the effect of using assay-specific upper limits of normal for AST, as reported by each centre. Finally, we assessed the effect of using a lower liver stiffness threshold to define cirrhosis (9.5 kPa).^13^

To explore an association with cirrhosis and significant fibrosis, we used mixed effects logistic regression models with study site-specific random effects. We explored variables that were anticipated a priori to be clinically important comprising age, sex, BMI category and reason for hepatitis B testing (asymptomatic vs suspected liver disease). To examine an association between APRI test sensitivity and liver stiffness measurement among patients with cirrhosis, we used the Wilcoxon rank sum test and plotted the distribution of liver stiffness and APRI classification using a kernel density plot and a restricted cubic spline plot for liver stiffness against test sensitivity. Analyses were conducted in R version 4.1.0 (R Foundation for Statistical Computing, Austria), JAGS 4.3.0, using the rjags package (v4.10)^22^ and Stata v17 (Statacorp, USA). All R and JAGS code is available from GitHub (https://github.com/gitMarcH/HEPSANET).

### Ethical review

The study protocol was registered (PROSPERO CRD42020218043). The study was reported in accordance with the PRISMA-IPD guidelines.^23^ Each participating centre obtained permission from local research ethical review committees (Appendix 6).

## RESULTS

Database searches identified 1,334 articles following removal of duplicates. After screening of title and abstract, the full text of 72 articles were obtained for detailed review (Appendix 7). In total 24 articles met our inclusion criteria. Cohorts represented by HEPSANET group members were described in 21 articles.^11,13,24-42^ We contacted corresponding authors from the remaining 3 articles,^43-45^ and all authors agreed to contribute individual patient data. We additionally included unpublished data from three sites (Cape Town, South Africa; Thiès, Senegal; and Dakar, Senegal) and data from one additional study published since the search was conducted from Malawi.^46^

Overall, contributing centres reported on 3960 HBsAg positive patients from 12 cohorts in eight countries comprising Burkina Faso, Ethiopia, The Gambia, Malawi, Nigeria, Senegal, South Africa, and Zambia (Figure 1). We excluded 411 ineligible participants who were HIV positive or with unknown HIV status, had received hepatitis B antiviral therapy, had positive anti-HCV or anti-HDV serology or had ALT/AST >5x the upper limit of normal (Figure 2). Characteristics of individual cohorts are reported in Appendix 3.

**Figure 1:**
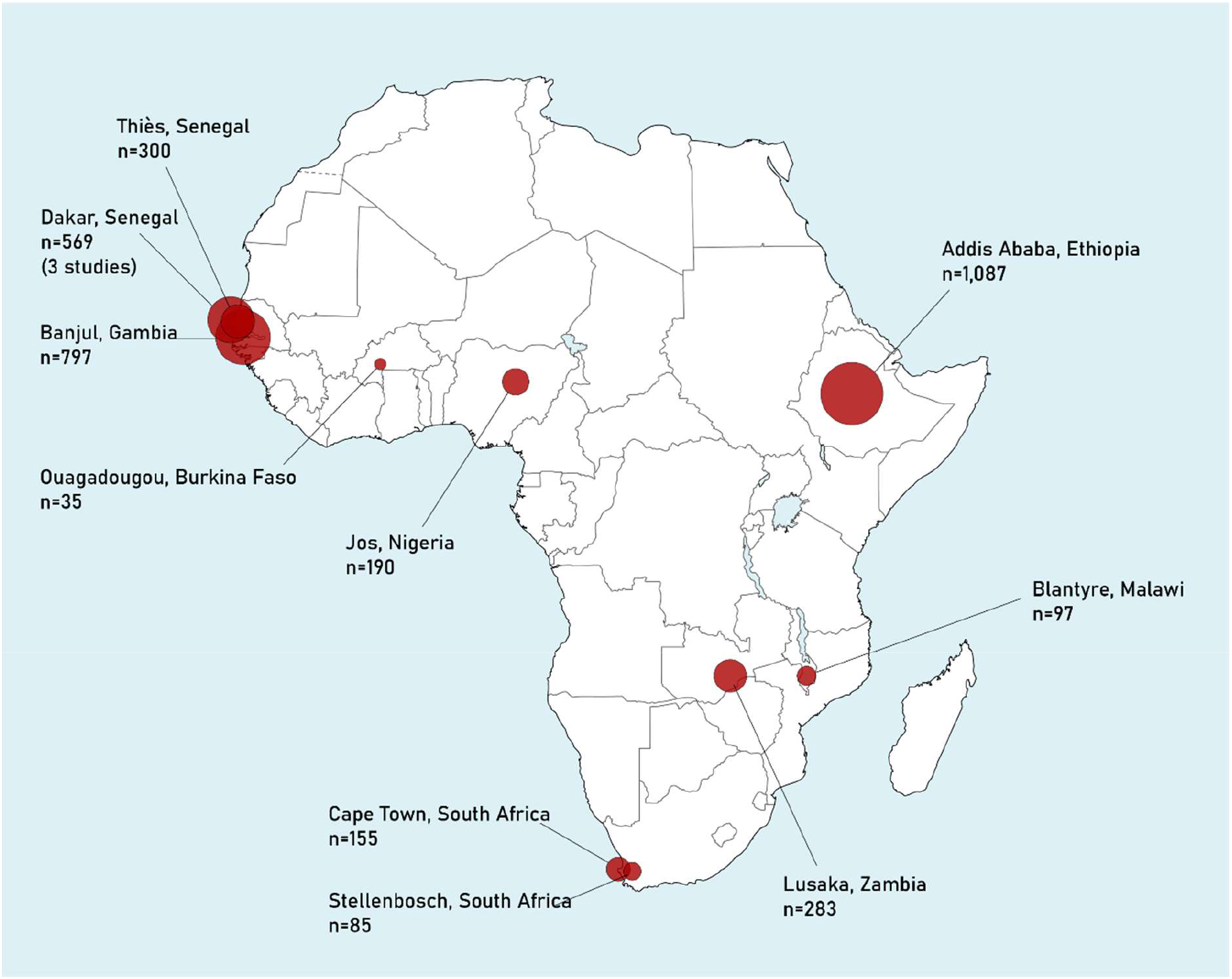
Map of included studies. Abbreviations: n; the number of hepatitis B patients included from each site in the current analysis. Area of circles is proportionate to cohort size.

**Figure 2:**
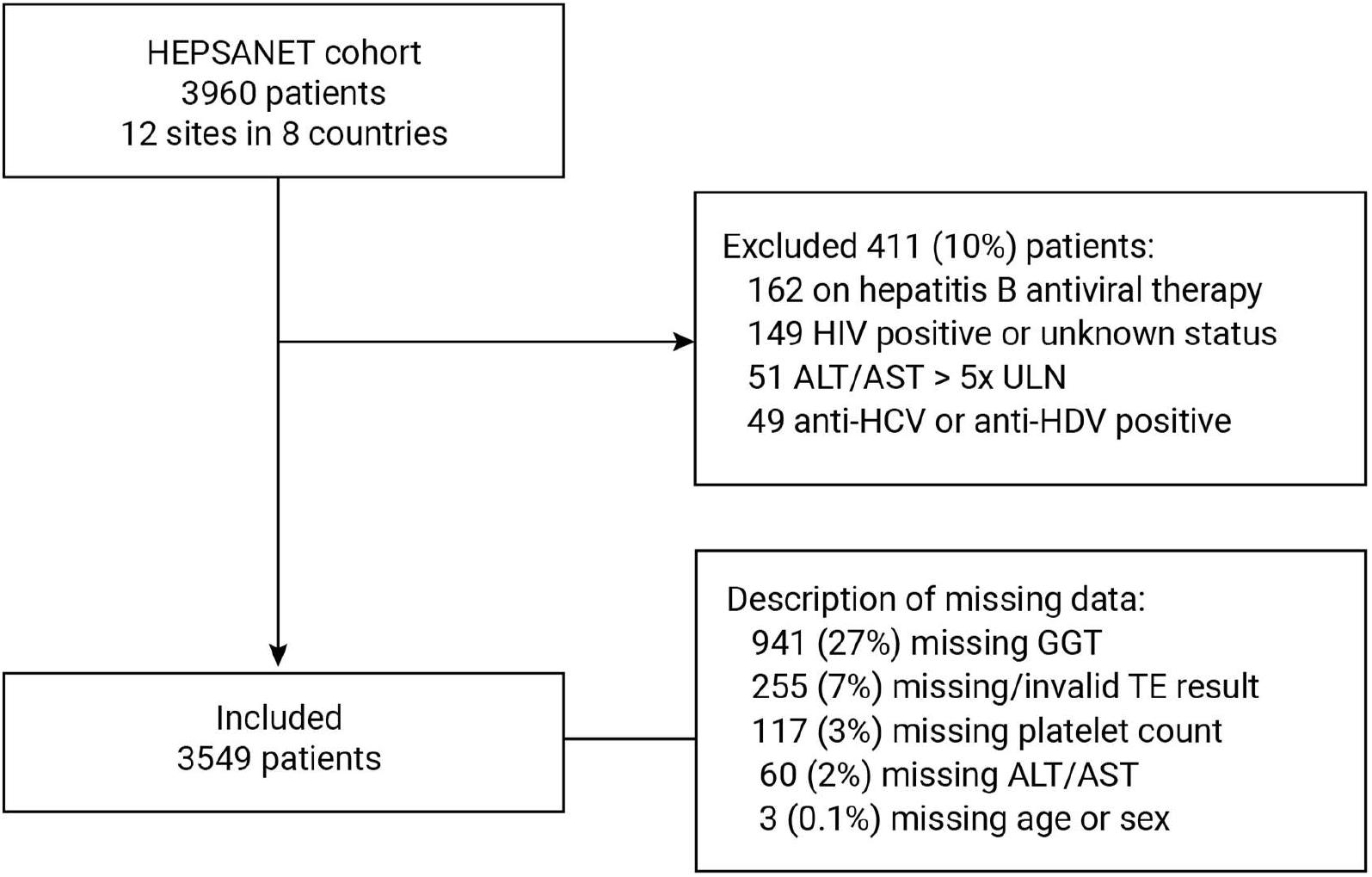
Flowchart of HEPSANET data sources.

Assessment of study quality indicated an overall low risk of bias according to QUADAS-2 criteria (Appendix 8). All studies avoided a case-control design and systematically performed both standardised index and reference tests in study participants. Two cohorts had specific criteria that could reduce applicability: one required patients were symptom-free and had HBV DNA >3.2 log10 IU/ml,^39^ and one excluded patients with BMI >28 kg/m^2^.^43^ Three studies performed TE in a subset of the overall cohort with non-random selection: two due to equipment availability,^12,37^ and one at clinician’s discretion without specifying criteria.^45^ Two studies had significant loss to follow up between community diagnosis and evaluation.^38,46^

Among 3,549 included participants, median age was 33 years (inter-quartile range (IQR) 28-41) and 60% were male (Table 1). Recruitment was hospital-based in three-quarters, with the remainder from community-based studies. A fifth of patients met BMI criteria for overweight and 7% were obese, whereas 5% reported hazardous alcohol consumption. In over 80% of cases (2,824/3,498), the reason for testing for hepatitis B was *asymptomatic screening*, as part of antenatal care, at blood donation, or because of a family contact with HBV, whereas for 674 individuals (19%) testing was performed due to *suspected liver disease, viz* due to symptoms or clinical signs, or abnormal liver function tests. The prevalence of cirrhosis was 7.4% overall: 2.5% among asymptomatically screened populations and 26.5% among patients with suspected liver disease. Cirrhosis was more prevalent among male (9.9%) relative to female participants (3.4%) and cirrhosis prevalence increased with age (Appendix 9). Significant fibrosis was observed in 17.5% overall; 11.5% among asymptomatic screening populations and 40.8% among patients with suspected liver disease. In a multivariable model, increased likelihood of significant fibrosis or cirrhosis was observed with increasing age, male gender, and suspected liver disease (relative to asymptomatic screening populations), whereas overweight or obese patients were less likely to have significant fibrosis or cirrhosis (Appendix 10). Ascites was observed in 90/3549 (2.5%) participants. Among patients with ascites with valid liver stiffness measurements (LSM), 78/88 (88.6%) had cirrhosis (F4), 8/88 (9.1%) had significant fibrosis (F2) and 2 (2.3%) had LSM within the normal range.

**Table 1:** Characteristics of included participants.

| Characteristic<br>n (%) or median (IQR) | Overall |  | Reason for HBV testing |  |  |  | Missing data<br>n (%) |  |
| --- | --- | --- | --- | --- | --- | --- | --- | --- |
|  |  |  | Asymptomatic<br>screening<br>populations<sup>a</sup> |  | Suspected liver<br>disease<sup>a</sup> |  |  |  |
| Total | 3,549 | (100) | 2,824 | (80.7) | 674 | (19.3) |  |  |
| Type of study |  |  |  |  |  |  | 0 | (0) |
| Hospital based | 2,648 | (74.6) | 1,923 | (68.1) | 674 | (100) |  |  |
| Community based | 901 | (25.4) | 901 | (31.9) | 0 | (0) |  |  |
| Country |  |  |  |  |  |  | 0 | (0) |
| Ethiopia | 1,038 | (29.3) | 717 | (25.4) | 321 | (47.6) |  |  |
| Senegal | 869 | (17.4) | 787 | (27.9) | 76 | (11.3) |  |  |
| The Gambia | 797 | (24.6) | 797 | (28.2) | 0 | (0) |  |  |
| Zambia | 283 | (8.0) | 256 | (9.1) | 18 | (2.7) |  |  |
| South Africa | 240 | (6.8) | 194 | (6.9) | 45 | (6.7) |  |  |
| Nigeria | 190 | (5.4) | 0 | (0) | 190 | (28.2) |  |  |
| Malawi | 97 | (2.7) | 73 | (2.6) | 24 | (3.6) |  |  |
| Burkina Faso | 35 | (1.0) |  |  |  |  |  |  |
| Sex, male | 2,134 | (60.2) | 1,656 | (58.7) | 450 | (66.8) | 1 | (0) |
| Age, years |  |  |  |  |  |  | 2 | (0.1) |
| Median (IQR) | 33 | (28, 41) | 33 | (28, 41) | 34 | (28, 42) |  |  |
| 14-29 | 1,147 | (32.3) | 919 | (32.6) | 208 | (30.9) |  |  |
| 30-39 | 1,345 | (37.9) | 1078 | (38.2) | 250 | (37.1) |  |  |
| 40-49 | 635 | (17.9) | 501 | (17.8) | 125 | (18.6) |  |  |
| 50-59 | 292 | (8.2) | 222 | (7.9) | 65 | (9.6) |  |  |
| 60-69 | 105 | (3.0) | 84 | (3.0) | 21 | (3.1) |  |  |
| ≥70 | 23 | (0.7) | 18 | (0.6) | 5 | (0.7) |  |  |
| Body mass index<br>(kg/m<sup>2</sup>) | 22.4 | (20.0, 25.5) | 22.5 | (20.0, 25.6) | 22.2 | (19.7, 24.7) | 426 | (12.0) |
| Overweight<br>(BMI 25.0-29.9) | 675 | (21.6) | 536 | (21.8) | 125 | (20.5) |  |  |
| Obese (BMI≥30.0) | 205 | (6.6) | 178 | (7.2) | 20 | (3.3) |  |  |
| Hazardous alcohol<br>consumption<br>reported<sup>b</sup> | 118 | (4.7) | 89 | (4.4) | 26 | (6.3) | 1,047 | (29.5) |
| HBeAg positive | 278 | (9.0) | 170 | (7.0) | 100 | (17.0) | 474 | (13.4) |
| HBV DNA (log<sub>10</sub> IU/ml),<br>median (IQR)<sup>c</sup> | 2.7 | (1.8, 3.7) | 2.6 | (1.7, 3.6) | 3.5 | (2.6, 5.9) | 577 | (16.3) |
| <2000 IU/ml | 1,920 | (64.6) | 1,712 | (67.8) | 182 | (44.7) |  |  |
| 2000-19,999 IU/ml | 541 | (18.2) | 461 | (18.3) | 70 | (17.2) |  |  |
| ≥20,000 IU/ml | 511 | (17.2) | 352 | (13.9) | 155 | (38.1) |  |  |
| Liver stiffness<br>measurement (kPa) |  |  |  |  |  |  | 255 | (7.9) |
| Median (IQR) | 5.5 | (4.5, 6.9) | 5.3 | (4.4, 6.6) | 6.8 | (5.0, 13.6) |  |  |
| ≤7.9 | 2,680 | (82.5) | 2,285 | (88.5) | 395 | (59.2) |  |  |
| 8.0-9.5 | 202 | (6.2) | 150 | (5.8) | 52 | (7.8) |  |  |
| 9.6-12.2 | 126 | (3.9) | 83 | (3.2) | 43 | (6.5) |  |  |
| >12.2 | 241 | (7.4) | 64 | (2.5) | 177 | (26.5) |  |  |

Abbreviations: ULN, Upper limit of normal; IQR, interquartile range.
<sup>a</sup>Reason for testing for hepatitis B was missing for 51 (1.4%) of participants. <sup>b</sup>Hazardous alcohol consumption was as defined by each centre; centre-specific definitions are reported in the supplementary appendix.

APRI and GPR had the best discriminant performance, with AUROC of 0.81 and 0.82 for cirrhosis and 0.75 and 0.76 for significant fibrosis, respectively. FIB-4 had relatively lower AUROC of 0.77 for cirrhosis and 0.68 for significant fibrosis. ALT as a standalone marker was associated with the lowest performance (Figure 3). Performance was significantly better for the diagnosis of cirrhosis relative to the diagnosis of significant fibrosis for all evaluated biomarkers (Figure 3).

**Figure 3:**
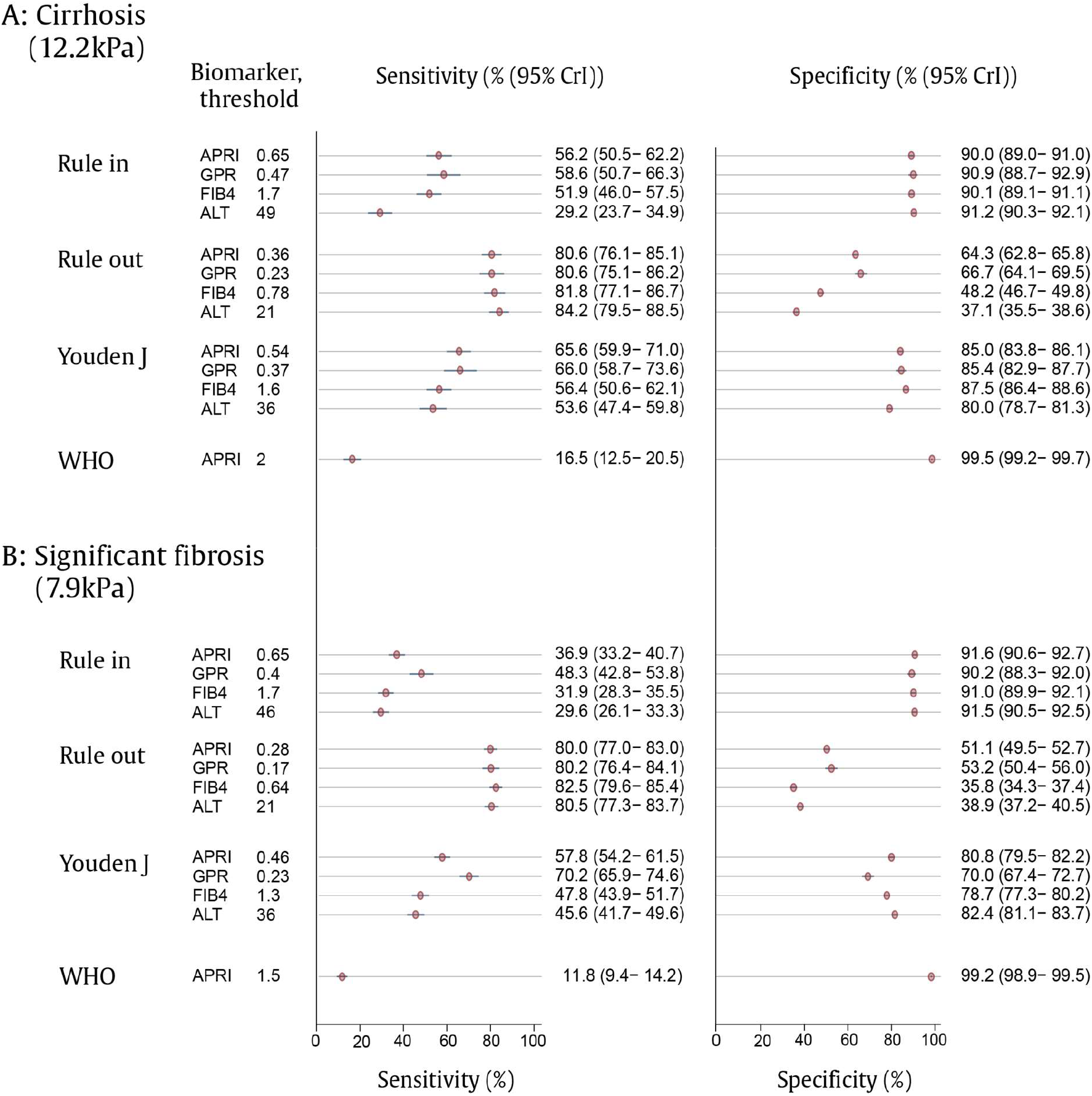
Performance of non-invasive biomarkers APRI, GPR, FIB-4 and ALT compared to a transient elastography reference standard for the diagnosis of A) cirrhosis and B) significant fibrosis: Bayesian random effects model^a^. Abbreviations: CrI, credible interval; APRI, asparatate aminotransferase-to-platelet ratio index; FIB-4, fibrosis-4 score; GPR, gamma-glutamyl transferase-to-platelet ratio; ALT, alanine aminotransferase. ^a^Bayesian bivariate random effects model adjusted for sex, study, hazardous alcohol consumption, reason for testing (suspected liver disease or asymptomatic screening) and categorical body mass index; summary statistics show population average. Youden’s J = maximisation of Youden’s J statistic, with equal weight to sensitivity and specificity (J = sensitivity + specificity - 1). Rule in models were chosen where specificity exceeds 90%; rule out models where sensitivity >80%.

We first evaluated the existing WHO recommendations to use APRI with a threshold of 2.0 for cirrhosis and 1.5 for significant fibrosis. These thresholds were associated with a sensitivity of 16.5% (95% credible interval (CrI) 12.5–20.5) for the diagnosis of cirrhosis and 11.8% (95% CrI 9.4–14.2) for significant fibrosis, whereas the specificity was 99.5% (95% CrI 99.2–99.7) for cirrhosis and 99.2% (95% CrI 98.9–99.5) for significant fibrosis.

We next developed new and optimised rule-out thresholds where sensitivity exceeded 80%, *viz* where 80% of patients with cirrhosis have a positive biomarker test (Figure 3). For APRI this threshold was 0.36 with a sensitivity of 80.6% (95% CrI 76.1–85.1) and a specificity of 64.3% (95% CrI 62.8–65.8); for GPR this was 0.23 with a sensitivity of 80.6% (95% CrI 75.1– 86.2) and a specificity of 66.7% (95% CrI 64.1–69.5).

Thirdly, we developed new and optimised rule-in thresholds where specificity exceeded 90%, *viz* where at least 90% of patients who did not have cirrhosis by the reference test had a negative biomarker test. For APRI the threshold 0.65 was associated with a specificity of 90.0% (95% CrI 89.0-91.0) and a sensitivity of 56.2% (95% CrI 50.5-62.2); for GPR the threshold was 0.47 with a specificity of 90.9% (95% CrI 88.7–92.9) and a sensitivity of 58.6 (95% CrI 50.7–66.3). We assessed the proposed cut-offs in each individual site (Appendix 11). Significant inter-site heterogeneity was observed; however, due to a small number of patients with cirrhosis at most sites, confidence intervals were wide at the level of individual centres.

Predictive values were strongly associated with disease prevalence (Table 2). Using models stratified by reason for testing, among asymptomatic screening populations, at the rule-in thresholds for APRI (0.65) and GPR (0.47), positive predictive values for cirrhosis were 17.0% for APRI and 20.1% for GPR, while negative predictive values were 98.4% and 98.5%, respectively. Among patients tested for suspected liver disease, the positive predictive values using the same thresholds were 70.5% and 74.9%, whereas negative predictive values were 77.3% and 73.0%, respectively. Significant trade-offs between optimising test sensitivity and specificity were noted with the use of all biomarkers. The implications of these trade-offs are illustrated in figures 4 and 5. Lower biomarker thresholds were associated with substantial over-diagnosis of cirrhosis (Figure 5).

**Table 2:** Association between predictive values and study populations for APRI and GPR.

| Population, diagnostic target<sup>a</sup> | Biomarker | Cut-off | PPV (%) | NPV (%) | Sensitivity (%) | Specificity (%) |
| --- | --- | --- | --- | --- | --- | --- |
| Asymptomatic screening, cirrhosis | APRI rule-in | 0.65 | 17.0 (13.9-20.2) | 98.4 (98.0-98.8) | 56.4 (45.9-66.9) | 90.6 (89.5-91.6) |
|  | GPR rule-in | 0.47 | 20.1 (16.1-24.3) | 98.5 (98.1-99.0) | 69.7 (48.0-71.4) | 91.9 (90.5-93.3) |
|  | APRI Youden's J | 0.54 | 14.0 (11.9-16.0) | 98.8 (98.4-99.2) | 68.8 (58.6-78.6) | 85.6 (84.3-86.8) |
|  | GPR Youden's J | 0.37 | 13.9 (11.3-16.5) | 98.6 (98.1-99.0) | 63.4 (51.9-74.5) | 86.7 (85.0-88.4) |
|  | APRI rule-out | 0.36 | 7.4 (6.7-8.1) | 99.0 (98.7-99.4) | 80.7 (73.7-87.8) | 65.5 (64.0-67.1) |
|  | GPR rule-out | 0.23 | 7.8 (6.8-8.8) | 98.9 (98.5-99.4) | 79.4 (69.5-88.5) | 68.2 (66.1-70.3) |
| Suspected liver disease, cirrhosis | APRI rule-in | 0.65 | 70.5 (65.4-75.6) | 77.3 (74.6-79.9) | 56.6 (49.9-62.8) | 86.2 (83.2-89.1) |
|  | GPR rule-in | 0.47 | 74.9 (48.9-93.7) | 73.0 (68.4-77.2) | 42.5 (32.0-52.5) | 90.6 (73.9-98.7) |
|  | APRI Youden's J | 0.54 | 66.0 (61.5-70.3) | 79.4 (76.6-82.3) | 64.0 (58.0-70.0) | 80.7 (77.4-84.0) |
|  | GPR Youden's J | 0.37 | 72.0 (51.7-87.8) | 76.8 (72.3-81.0) | 55.2 (44.2-64.9) | 86.3 (70.0-96.3) |
|  | APRI rule-out | 0.36 | 51.9 (49.0-54.7) | 83.2 (79.3-87.0) | 80.3 (75.1-85.4) | 56.7 (52.4-60.6) |
|  | GPR rule-out | 0.23 | 49.7 (40.8-63.7) | 81.2 (73.5-88.6) | 80.2 (73.4-86.6) | 51.3 (34.4-73.9) |
| Asymptomatic screening, significant fibrosis | APRI rule-in | 0.65 | 10.7 (8.5-12.9) | 97.4 (97.3-97.6) | 28.8 (23.8-33.8) | 91.8 (90.7-92.9) |
|  | GPR rule-in | 0.40 | 12.2 (9.8-14.6) | 97.7 (97.5-98.0) | 38.5 (32.1-44.8) | 90.5 (89.0-92.0) |
|  | APRI Youden's J | 0.46 | 8.0 (7.0-9.0) | 97.9 (97.7-98.1) | 48.8 (43.7-53.9) | 80.9 (79.4-82.3) |
|  | GPR Youden's J | 0.23 | 7.0 (6.3-7.7) | 98.3 (98.0-98.6) | 63.9 (58.1-69.7) | 71.0 (68.8-73.2) |
|  | APRI rule-out | 0.27 | 4.7 (4.4-5.0) | 98.5 (98.2-98.7) | 78.9 (75.0-82.9) | 45.4 (43.6-47.1) |
|  | GPR rule-out | 0.17 | 5.4 (5.0-5.8) | 98.5 (98.2-98.8) | 76.5 (71.4-81.1) | 54.1 (51.7-56.4) |
| Suspected liver disease, significant fibrosis | APRI rule-in | 0.65 | 75.1 (68.8-80.7) | 74.9 (72.9-76.9) | 47.6 (42.3-53.2) | 90.8 (88.0-93.4) |
|  | GPR rule-in | 0.40 | 83.6 (69.3-93.2) | 79.6 (74.3-83.5) | 59.0 (46.0-68.4) | 93.1 (85.5-97.6) |
|  | APRI Youden's J | 0.46 | 66.9 (62.5-71.5) | 81.6 (79.2-84.1) | 69.0 (64.1-73.7) | 80.1 (76.2-83.8) |
|  | GPR Youden's J | 0.23 | 60.2 (48.5-70.6) | 83.4 (78.6-88.1) | 76.2 (67.7-83.5) | 69.9 (53.6-82.8) |
|  | APRI rule-out | 0.27 | 44.7 (42.5-47.0) | 83.8 (79.4-88.4) | 87.9 (84.2-91.4) | 36.6 (31.8-41.8) |
|  | GPR rule-out | 0.17 | 51.8 (43.8-60.3) | 84.4 (78.1-89.8) | 82.8 (74.0-90.0) | 54.5 (38.4-69.9) |

<sup>a</sup>Cirrhosis prevalence was 2.5% among people tested for hepatitis B as part of asymptomatic screening, and 26.5% among people tested for hepatitis B due to suspected liver disease (viz with symptoms, clinical signs, or abnormal liver function tests).

**Figure 4:**
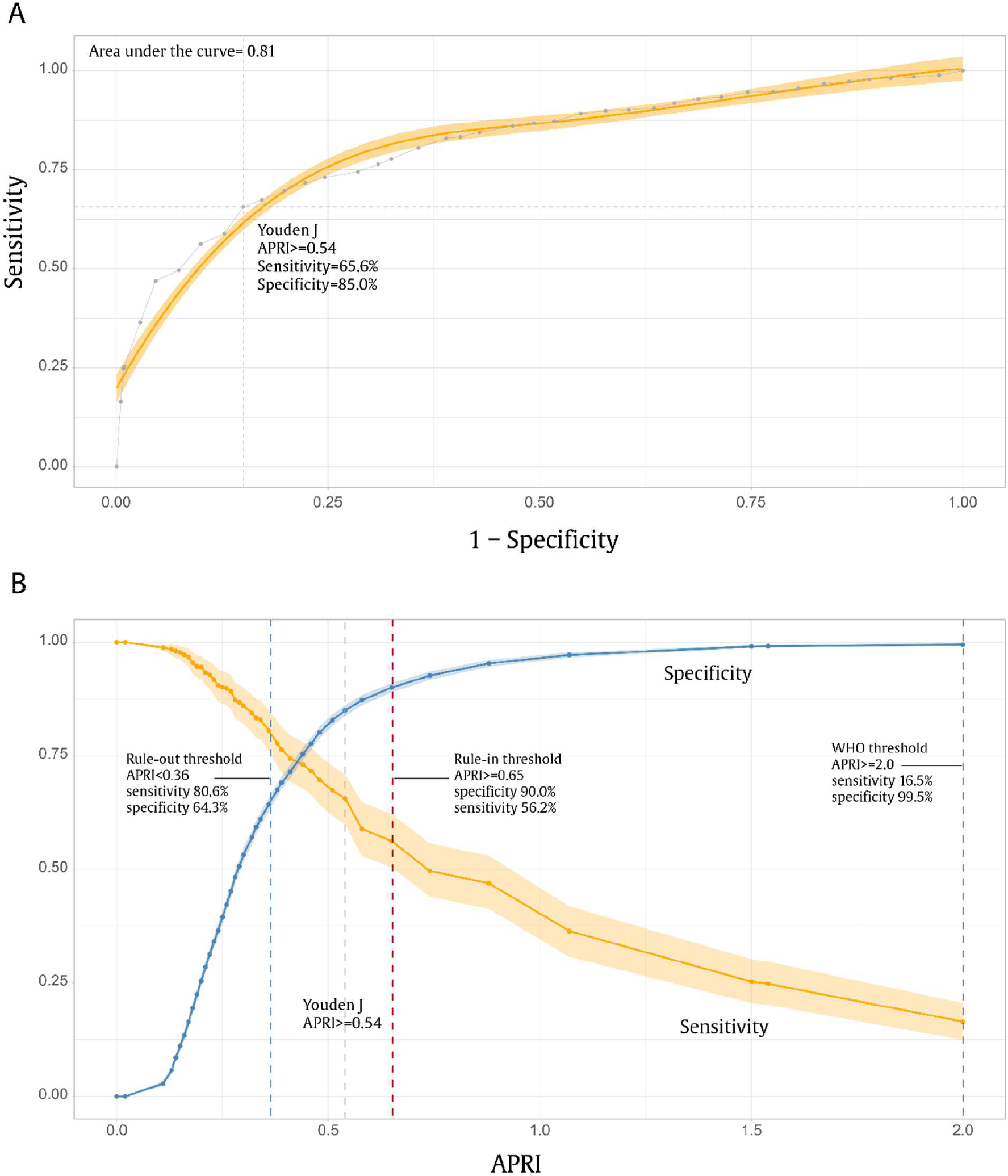
Relationship between sensitivity and specificity for APRI used to diagnosis cirrhosis (12.2 kPa): A) Receiver operating curve; B) Sensitivity and specificity as a function of APRI^a^. ^a^ Bayesian bivariate random effects model fitted for different thresholds of APRI using 60 equally spaced quantiles. The ROC curve (part A, shaded orange) is a shape-constrained generalized additive model fitted to the raw estimates. Area under the curve is computed from the raw estimates. Shaded areas surrounding lines represent 95% credible intervals. Specificity is defined as the probability of a negative test given the absence of cirrhosis; sensitivity is defined as the probability of a positive test, given the presence of cirrhosis.

**Figure 5:**
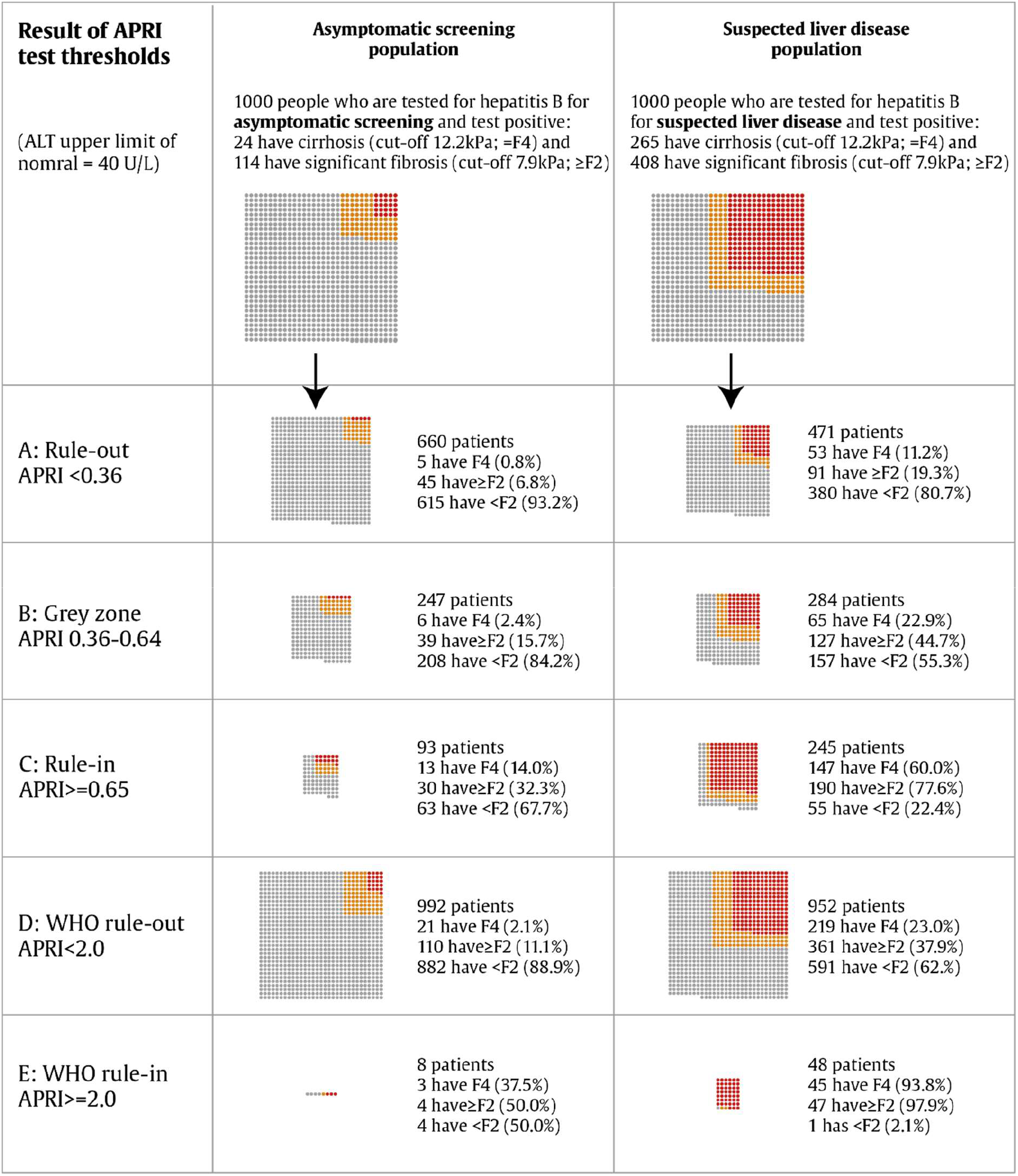
APRI biomarker testing in asymptomatic vs. symptomatic populations using rule-in and rule-out thresholds (A-C) and the WHO recommended threshold (D-E)^a^. ^a^ A single grey dot represents one person without significant fibrosis (F0-1), one orange dot represents a person with significant fibrosis (F2-3) and one red dot represents a person with cirrhosis (F4). Individual proportions rounded to nearest whole individual and represent the effect of applying APRI thresholds to the average populations included in HEPSANET cohorts, stratified by reason for hepatitis B testing.

Patient level co-variates significantly associated with model performance at the APRI and GPR rule-in thresholds included hazardous alcohol consumption, suspected liver disease as the reason for testing, and female sex (Appendix 12). Hazardous alcohol consumption reduced test specificity for both biomarkers. Specificity was improved for APRI and GPR for female relative to male participants with no significant effect of sex on sensitivity. For the GPR model, being overweight reduced test specificity.

We evaluated the model with several sensitivity analyses. First, we assessed the effect of excluding patients for whom ascites was observed, since the probability of cirrhosis was very high among this group; this exclusion had a limited effect on diagnostic performance (Appendix 13). Secondly, we observed the effect of using sex-specific ULN for AST (male 30 U/L; female 19 U/L). This was associated with reduced sensitivity and discriminant performance relative to a fixed ULN of 40 U/L for all participants. Thirdly, we assessed the effect of using centre-specific ULN thresholds; these ranged from 25 to 60 U/L for AST and 24 to 85 U/L for GGT. This was also associated with reduced diagnostic performance. Finally, we considered the effect of using a lower TE value of 9.5 kPa for the diagnosis of cirrhosis. At this lower threshold, relative to 12.2 kPa, for both APRI and GPR, test performance was poorer, with lower sensitivity and specificity at rule-in and rule-out thresholds (Appendix 13).

In an exploratory analysis, we compared patients with cirrhosis who had false negative APRI classification results with those correctly classified to identify factors associated with impaired test sensitivity. The lowest sensitivity for APRI was observed among patients with LSM just above the threshold (12.2 kPa) for cirrhosis, and sensitivity increased with increasing liver stiffness with the effect plateauing above 30 kPa (Appendix 14).

## DISCUSSION

In this collaborative study of over 3500 patients with CHB in sSA, we found that cirrhosis was present in 7% and significant fibrosis in 18% of patients at pre-therapy evaluation, respectively. The reason for testing for hepatitis B was strongly associated with the prevalence of liver disease, with a cirrhosis prevalence of 3% among asymptomatic screening populations and 27% among patients tested due to suspected liver disease.

We evaluated the diagnostic performance of the liver fibrosis biomarker APRI, recommended in the first WHO CHB treatment guidelines from 2015, which have been widely adopted into national programmes in sSA.^8^ Our analysis included all identified existing published data, together with novel unpublished data, offering the most comprehensive evaluation of these tools from the region to date. We found that the sensitivity of APRI to diagnose cirrhosis at the WHO-recommended threshold of 2.0 was only 16.5%, which confirms previous reports of low sensitivity in the region.^11-13^ We also identified new rule-in and rule-out thresholds suitable for sSA for APRI and GPR. The best test characteristics were observed for APRI and GPR for the detection of cirrhosis; an APRI rule-in threshold of 0.65 yielded 90.0% specificity and 56.2% sensitivity, whereas a rule-out threshold of 0.36 yielded 80.6% sensitivity and 64.3% specificity.

Our findings compare well with a recent large IPD analysis of mainly Asian patients using liver biopsy as a reference standard.^47^ The authors found that the conventional APRI and FIB-4 thresholds were unsuitable for patient management and identified new rule-out thresholds for both APRI (0.45) and FIB-4 (0.70) that were significantly lower, in line with our findings. Of note, both our study and the study by Sonneveld and colleagues, applied a fixed ULN for AST. In a sensitivity analysis, we observed a poorer performance of APRI when using assay-specific or sex-specific ULN. It is likely that the variable performance of APRI in previous studies could be, at least partly, explained by the differing definitions of ULN for AST.

The new thresholds identified in our study for APRI and GPR were particularly suitable at *ruling out* disease (i.e. they had high negative predictive values). The ability to *rule in* disease (i.e. positive predictive value) was much poorer. This phenomenon reflects a common problem with screening tests, particularly in a community setting, resulting in a significant rate of overdiagnosis when the prevalence and pre-test probability of disease is low.^48^ One might argue that some degree of overdiagnosis of cirrhosis is acceptable since many of those with a high APRI score (in the absence of cirrhosis) have F2/F3 fibrosis or active hepatic inflammation and need antiviral therapy to prevent progression to cirrhosis or HCC. Overdiagnosis would lead to wider use of nucleoside analogues, the preferred treatment for CHB, which are safe and well-tolerated, and offer the benefit of pre-exposure prophylaxis for HIV, of particular importance in high prevalence settings.^49,50^ On the other hand, excessive overtreatment could overstretch health budgets. In future work within the HEPSANET collaboration we will aim to optimise treatment eligibility criteria for sSA, beyond the present question of fibrosis assessment.

Our findings will stimulate debate on several alternative responses. These include arguments for a universal test-and-treat strategy, or to apply a significantly lower eligibility threshold for antiviral treatment, or to use the biomarkers with a rule-out approach aiming to discharge from follow-up those at low risk of disease progression. Accurately identifying low-risk patients who can be discharged from follow-up is a particularly important priority in high-prevalence, resource-limited settings in sSA, and will require prospective data. We aim to perform a cost-effectiveness analysis within the HEPSANET collaboration to shed light on the balance between over-treatment and widening access to care. The optimal strategy is likely to vary in different settings depending on disease prevalence and available resources.

The intended use of these biomarkers should be kept in mind when interpreting our results. Importantly, the result of a fibrosis marker is never the only criterion to assess whether a patient needs antiviral therapy. Clinical assessment, ALT, age, and family history are available in most settings, and HBeAg and HBV DNA might be available at larger centres in low- and middle-income countries. Thus, a full patient assessment will perform better than an evaluation considering APRI or GPR alone, as presented in this study.

This study had some limitations. First, it is an analysis of heterogenous populations. Accordingly significant heterogeneity was observed between centres that was only partly explained by the examined participant characteristics. Second, only untreated CHB patients were included in the analysis and the results might not apply to those who have commenced antiviral therapy. Third, APRI and GPR displayed reduced specificity in patients with alcohol abuse. It is well-known that alcohol may cause elevations of both AST and GGT and reductions in platelet count;^51^ hence, fibrosis scores based on AST and GGT must be used with caution in patients with known alcohol misuse. Finally, although there was an overall low risk of bias associated with the studies, a small number had non-random selection criteria or specific exclusions that could limit applicability.

The study has significant strengths. This is the largest and most comprehensive analysis from the region, including all previously published individual patient data and including previously unpublished data from newer cohorts. Our collaborative network has a good geographical representation, with studies from east, west and southern Africa. We therefore believe our results are generalisable. The bivariate random effects meta-analysis (BRMA) modelling framework we used has been widely advocated for, in diagnostic accuracy meta-analysis.^21,52^ The BRMA framework accounts for both within study precision of estimates and heterogeneity between studies. The model allowed us to model the effect of patient-level covariates on diagnostic accuracy. Further, the models jointly model sensitivity and specificity, capturing the dependence between these two parameters. Finally, the Bayesian estimation framework we used allows a principled treatment of missing data.

In conclusion, APRI had a poor sensitivity for cirrhosis at the WHO-recommended threshold of 2.0 in sSA. We developed new rule-in and rule-out thresholds for APRI and GPR which had better discriminatory properties for the detection of cirrhosis. The non-invasive biomarkers were better at ruling out than ruling in disease, and when applied as screening tools on a general population a certain degree of over-diagnosis must be assumed. Our data are important for informing practice in sSA and should be considered in the next revision of the WHO hepatitis B guidelines. Our findings highlight the need for new biomarkers, expanded access to treatment, and new ways of thinking about treatment eligibility for CHB in sSA.

## Data Availability

All data produced in the present study are available upon reasonable request to the authors

## ACKNOWLEDGEMENTS

We would like to thank the staff at participating centres in the HEPSANET collaboration who contributed to patient follow-and data collection. The authors assume full responsibility for analyses and interpretation of these data.

## REFERENCES

1. World Health Organisation. Global progress report on HIV, viral hepatitis and sexually transmitted infections, 2021. Geneva, Switzerland 2021.

2. Trepo C, Chan HL, Lok A. Hepatitis B virus infection. Lancet. 2014;384(9959):2053–2063.

3. Kim WR, Loomba R, Berg T, et al. Impact of long-term tenofovir disoproxil fumarate on incidence of hepatocellular carcinoma in patients with chronic hepatitis B. Cancer. 2015;121(20):3631–3638.

4. Marcellin P, Gane E, Buti M, et al. Regression of cirrhosis during treatment with tenofovir disoproxil fumarate for chronic hepatitis B: a 5-year open-label follow-up study. Lancet. 2013;381(9865):468–475.

5. European Association for the Study of the Liver. EASL 2017 Clinical Practice Guidelines on the management of hepatitis B virus infection. J Hepatol. 2017;67(2):370–398.

6. Sarin SK, Kumar M, Lau GK, et al. Asian-Pacific clinical practice guidelines on the management of hepatitis B: a 2015 update. Hepatol Int. 2016;10(1):1–98.

7. Terrault NA, Lok ASF, McMahon BJ, et al. Update on prevention, diagnosis, and treatment of chronic hepatitis B: AASLD 2018 hepatitis B guidance. Hepatology. 2018;67(4):1560–1599.

8. World Health Organisation. Guidelines for the Prevention, Care and Treatment of Persons with Chronic Hepatitis B Infection. In: Geneva: World Health Organization; 2015.

9. Wai CT, Greenson JK, Fontana RJ, et al. A simple noninvasive index can predict both significant fibrosis and cirrhosis in patients with chronic hepatitis C. Hepatology. 2003;38(2):518–526.

10. Sterling RK, Lissen E, Clumeck N, et al. Development of a simple noninvasive index to predict significant fibrosis in patients with HIV/HCV coinfection. Hepatology. 2006;43(6):1317–1325.

11. Desalegn H, Aberra H, Berhe N, Gundersen SG, Johannessen A. Are non-invasive fibrosis markers for chronic hepatitis B reliable in sub-Saharan Africa? Liver Int. 2017;37(10):1461–1467.

12. Bonnard P, Sombie R, Lescure FX, et al. Comparison of elastography, serum marker scores, and histology for the assessment of liver fibrosis in hepatitis B virus (HBV)-infected patients in Burkina Faso. Am J Trop Med Hyg. 2010;82(3):454–458.

13. Lemoine M, Shimakawa Y, Nayagam S, et al. The gamma-glutamyl transpeptidase to platelet ratio (GPR) predicts significant liver fibrosis and cirrhosis in patients with chronic HBV infection in West Africa. Gut. 2016;65(8):1369–1376.

14. Spearman CW, Afihene M, Ally R, et al. Hepatitis B in sub-Saharan Africa: strategies to achieve the 2030 elimination targets. Lancet Gastroenterol Hepatol. 2017;2(12):900–909.

15. Singh S, Fujii LL, Murad MH, et al. Liver stiffness is associated with risk of decompensation, liver cancer, and death in patients with chronic liver diseases: a systematic review and meta-analysis. Clin Gastroenterol Hepatol. 2013;11(12):1573–1584.e1571-1572; quiz e1588-1579.

16. Li Y, Huang YS, Wang ZZ, et al. Systematic review with meta-analysis: the diagnostic accuracy of transient elastography for the staging of liver fibrosis in patients with chronic hepatitis B. Alimentary pharmacology & therapeutics. 2016;43(4):458–469.

17. Mehta SH, Lau B, Afdhal NH, Thomas DL. Exceeding the limits of liver histology markers. J Hepatol. 2009;50(1):36–41.

18. Whiting PF, Rutjes AW, Westwood ME, et al. QUADAS-2: a revised tool for the quality assessment of diagnostic accuracy studies. Annals of internal medicine. 2011;155(8):529–536.

19. EASL-ALEH Clinical Practice Guidelines: Non-invasive tests for evaluation of liver disease severity and prognosis. J Hepatol. 2015;63(1):237–264.

20. Saunders JB, Aasland OG, Babor TF, De la Fuente JR, Grant M. Development of the alcohol use disorders identification test (AUDIT): WHO collaborative project on early detection of persons with harmful alcohol consumption-II. Addiction. 1993;88(6):791–804.

21. Riley RD, Dodd SR, Craig JV, Thompson JR, Williamson PR. Meta-analysis of diagnostic test studies using individual patient data and aggregate data. Stat Med. 2008;27(29):6111–6136.

22. Plummer M. rjags: Bayesian graphical models using MCMC. R package version. 2019;4(10).

23. Stewart LA, Clarke M, Rovers M, et al. Preferred Reporting Items for a Systematic Review and Meta-analysis of Individual Participant Data: The PRISMA-IPD Statement. JAMA. 2015;313(16):1657–1665.

24. Aberra H, Desalegn H, Berhe N, et al. Early experiences from one of the first treatment programs for chronic hepatitis B in sub-Saharan Africa. BMC Infect Dis. 2017;17(1):438.

25. Aberra H, Desalegn H, Berhe N, et al. The WHO guidelines for chronic hepatitis B fail to detect half of the patients in need of treatment in Ethiopia. J Hepatol. 2019;70(6):1065–1071.

26. Cohen D, Ghosh S, Shimakawa Y, et al. Hepatitis B virus preS2Δ38-55 variants: A newly identified risk factor for hepatocellular carcinoma. JHEP Rep. 2020;2(5):100144.

27. Desalegn H, Aberra H, Berhe N, et al. Predictors of mortality in patients under treatment for chronic hepatitis B in Ethiopia: a prospective cohort study. BMC Gastroenterol. 2019;19(1):74.

28. Desalegn H, Aberra H, Berhe N, et al. Treatment of chronic hepatitis B in sub-Saharan Africa: 1-year results of a pilot program in Ethiopia. BMC Med. 2018;16(1):234.

29. Johannessen A, Aberra H, Desalegn H, Gordien E, Berhe N. A novel score to select patients for treatment in chronic hepatitis B: Results from a large Ethiopian cohort. J Hepatol. 2019;71(4):840–841.

30. Lemoine M, Shimakawa Y, Njie R, et al. Food intake increases liver stiffness measurements and hampers reliable values in patients with chronic hepatitis B and healthy controls: the PROLIFICA experience in The Gambia. Aliment Pharmacol Ther. 2014;39(2):188–196.

31. Lemoine M, Shimakawa Y, Njie R, et al. Acceptability and feasibility of a screen-and-treat programme for hepatitis B virus infection in The Gambia: the Prevention of Liver Fibrosis and Cancer in Africa (PROLIFICA) study. Lancet Glob Health. 2016;4(8):e559–567.

32. Nsokolo B, Kanunga A, Sinkala E, et al. Stage of disease in hepatitis B virus infection in Zambian adults is associated with large cell change but not well defined using classic biomarkers. Trans R Soc Trop Med Hyg. 2017;111(9):425–432.

33. Shimakawa Y, Boucheron P, Luong Nguyen LB, Lemoine M, Sombié R. Performance of two simplified HBV treatment criteria (TREAT-B score and WHO guidelines) in Burkina Faso, West Africa. J Hepatol. 2019;71(4):842–844.

34. Shimakawa Y, Lemoine M, Njai HF, et al. Natural history of chronic HBV infection in West Africa: a longitudinal population-based study from The Gambia. Gut. 2016;65(12):2007–2016.

35. Shimakawa Y, Ndow G, Njie R, et al. Hepatitis B Core-related Antigen: An Alternative to Hepatitis B Virus DNA to Assess Treatment Eligibility in Africa. Clin Infect Dis. 2020;70(7):1442–1452.

36. Shimakawa Y, Njie R, Ndow G, et al. Development of a simple score based on HBeAg and ALT for selecting patients for HBV treatment in Africa. J Hepatol. 2018;69(4):776–784.

37. Vinikoor MJ, Sinkala E, Kanunga A, et al. Chronic hepatitis B virus monoinfection at a university hospital in Zambia. World J Hepatol. 2018;10(9):622–628.

38. Vinikoor MJ, Sinkala E, Kanunga A, et al. Eligibility for hepatitis B antiviral therapy among adults in the general population in Zambia. PLoS One. 2020;15(1):e0227041.

39. Mbaye PS, Sarr A, Sire JM, et al. Liver stiffness measurement and biochemical markers in Senegalese chronic hepatitis B patients with normal ALT and high viral load. PLoS One. 2011;6(7):e22291.

40. Diallo I, Mbaye PS, Vray M, et al. Evaluation of fibrosis in chronic hepatitis B: comparison between liver biopsy and elastography (Fibroscanï¿½). J Africain Hepato-Gastroenterologie. 2016;10(3):132–137.

41. Diallo I, Ndiaye B, Fall CA, et al. Inactive hepatitis B carriers: outcomes of patients followed at Hôpital Principal de Dakar, Senegal. Pan Afr Med J. 2018;31:49.

42. Bonnard P, Sombié R, Lescure FX, et al. Comparison of elastography, serum marker scores, and histology for the assessment of liver fibrosis in hepatitis B virus (HBV)-infected patients in Burkina Faso. Am J Trop Med Hyg. 2010;82(3):454–458.

43. Ameh OA, Davwar PM, Duguru MJ, et al. Use of fibroscan in assessment of hepatic fibrosis in patients with chronic hepatitis B infection. Jos Journal of Medicine. 2018;12(2):9–14.

44. Maponga TG, Andersson MI, van Rensburg CJ, et al. HBV and HIV viral load but not microbial translocation or immune activation are associated with liver fibrosis among patients in South Africa. BMC Infect Dis. 2018;18(1):214.

45. Maponga TG, McNaughton AL, van Schalkwyk M, et al. Treatment advantage in HBV/HIV coinfection compared to HBV monoinfection in a South African cohort. J Infect. 2020;81(1):121–130.

46. Stockdale AJ, Meiring JE, Shawa IT, et al. Hepatitis B vaccination impact and the unmet need for antiviral treatment in Blantyre, Malawi. J Infect Dis. 2021.

47. Sonneveld MJ, Brouwer WP, Chan HL, et al. Optimisation of the use of APRI and FIB-4 to rule out cirrhosis in patients with chronic hepatitis B: results from the SONIC-B study. Lancet Gastroenterol Hepatol. 2019;4(7):538–544.

48. Manski CF. Bounding the Accuracy of Diagnostic Tests, With Application to COVID-19 Antibody Tests. Epidemiology. 2021;32(2):162–167.

49. Marcellin P, Wong DK, Sievert W, et al. Ten-year efficacy and safety of tenofovir disoproxil fumarate treatment for chronic hepatitis B virus infection. Liver Int. 2019;39(10):1868–1875.

50. Fonner VA, Dalglish SL, Kennedy CE, et al. Effectiveness and safety of oral HIV preexposure prophylaxis for all populations. AIDS (London, England). 2016;30(12):1973.

51. Gelsi E, Dainese R, Truchi R, et al. Effect of detoxification on liver stiffness assessed by Fibroscan® in alcoholic patients. Alcohol Clin Exp Res. 2011;35(3):566–570.

52. Reitsma JB, Glas AS, Rutjes AWS, Scholten RJPM, Bossuyt PM, Zwinderman AH. Bivariate analysis of sensitivity and specificity produces informative summary measures in diagnostic reviews. Journal of Clinical Epidemiology. 2005;58(10):982–990.

53. Stockdale AJ, Meiring JE, Shawa IT, et al. Hepatitis B Vaccination Impact and the Unmet Need for Antiviral Treatment in Blantyre, Malawi. The Journal of Infectious Diseases. 2021.

