## Appendices for "Diagnostic performance of non-invasive fibrosis markers for chronic hepatitis B in sub-Saharan Africa: a Bayesian individual patient data meta-analysis"

Appendix 1: Search methodology
Search date: 6th October 2020, no language or publication date restrictions applied

**PUBMED** (https://pubmed.ncbi.nlm.nih.gov/)- **697 results**

| |
|---|
| ("liver cirrhosis"[MeSH] OR "elasticity imaging techniques"[MeSH] OR fibrosis[tiab] OR cirrhosis[tiab] OR elastograph*[tiab] OR fibroscan[tiab] OR biopsy, needle[MeSH] OR "liver biops*"[tiab] OR metavir[tiab]) |
| AND (hepatitis B[MeSH] OR hepatitis b[tiab] OR HBV[tiab] OR HBsAg[tiab]) |
| AND (Africa[MeSH] OR Africa*[tiab] OR Angola[tiab] OR Benin[tiab] OR Botswana[tiab] OR "Burkina Faso"[tiab] OR Burundi[tiab] OR Cameroon[tiab] OR "Cape Verde"[tiab] OR "Central African Republic"[tiab] OR Chad[tiab] OR Comoros[tiab] OR Congo[tiab] OR Djibouti[tiab] OR "Equatorial Guinea"[tiab] OR Eritrea[tiab] OR Ethiopia[tiab] OR Gabon[tiab] OR Gambia[tiab] OR Ghana[tiab] OR Guinea[tiab] OR "Guinea Bissau"[tiab] OR "Ivory Coast"[tiab] OR "Cote d'Ivoire"[tiab] OR Kenya[tiab] OR Lesotho[tiab] OR Liberia[tiab] OR Madagascar[tiab] OR Malawi[tiab] OR Mali[tiab] OR Mauritania[tiab] OR Mauritius[tiab] OR Mozambique[tiab] OR Mocambique[tiab] OR Namibia[tiab] OR Niger[tiab] OR Nigeria[tiab] OR Principe[tiab] OR Reunion[tiab] OR Rwanda[tiab] OR "Sao Tome"[tiab] OR Senegal[tiab] OR Seychelles[tiab] OR "Sierra Leone"[tiab] OR Somalia[tiab] OR "South Africa"[tiab] OR Sudan[tiab] OR Swaziland[tiab] OR Tanzania[tiab] OR Togo[tiab] OR Tunisia[tiab] OR Uganda[tiab] OR Zambia[tiab] OR Zimbabwe[tiab]) |

**SCOPUS** (https://www.scopus.com/search/form.uri )- **947 results**

| |
|---|
| ( TITLE-ABS-KEY ( africa* OR angola OR benin OR botswana OR "Burkina Faso" OR burundi OR cameroon OR "Cape Verde" OR "Central African Republic" OR chad OR comoros OR congo OR djibouti OR "Equatorial Guinea" OR eritrea OR ethiopia OR gabon OR gambia OR ghana OR guinea OR "Guinea Bissau" OR "Ivory Coast" OR "Cote d'Ivoire" OR kenya OR lesotho OR liberia OR madagascar OR malawi OR mali OR mauritania OR mauritius OR mozambique OR mocambique OR namibia OR niger OR nigeria OR principe OR reunion OR rwanda OR "Sao Tome" OR senegal OR seychelles OR "Sierra Leone" OR somalia OR "South Africa" OR sudan OR swaziland OR tanzania OR togo OR tunisia OR uganda OR zambia OR zimbabwe) ) |
| AND ( TITLE-ABS-KEY ( "elasticity imaging" OR elastograph* OR fibroscan OR "needle biopsy" OR "liver biops*" OR metavir OR cirrhosis OR fibrosis ) ) |
| AND ( TITLE-ABS-KEY ( "hepatitis b" OR hbv OR hbsag ) ) |

**Africa Index Medicus (**https://indexmedicus.afro.who.int/**) - 11 results**

| |
|---|
| (tw:("elasticity imaging" OR elastograph* OR fibroscan OR "needle biopsy" OR "liver biops*" OR metavir OR cirrhosis OR fibrosis)) AND (tw:("hepatitis b" OR hbv OR hbsag)) |

**Africa Journals Online** (https://www.ajol.info/index.php/ajol) - **260 results**
Searched using Google Scholar (https://scholar.google.com/)

| |
|---|
| site:ajol.info (elastography OR liver biopsy OR fibroscan) AND "hepatitis B" |

Appendix 2: List of variables reported by HEPSANET participating sites

**2.1 Centre-specific variables**

| **Variable** | **Description/ criteria** |
|---|---|
| Country/ locale | Facility location |
| Study design | Community or hospital based |
| Criteria used for valid Fibroscan result | Centre definition |
| HBV DNA platform | Details of assay, manufacturer, platform |
| Biochemistry platform | Details of assay, manufacturer, platform for liver enzyme quantification |
| Schistosomiasis epidemiology | Describe whether endemic hepatic schistosomiasis (*S. mansoni)* |
| Schistosomiasis diagnosis | Method of diagnostic evaluation for schistosomiasis among centres with endemic disease |
| Harmful alcohol definition | Definition used for harmful alcohol consumption |

**2.2 Patient-specific variables (essential variables highlighted in bold)**

| **Variable** | **Description/ criteria** |
|---|---|
| **Patient age** | Unit: years |
| **Sex** | Male/female |
| **Pregnancy** | Current pregnancy |
| **Transient elastography** | Fasting (>2 hours) transient elastography result<br>Unit: kPa |
| **Alanine aminotransferase (ALT)** | Unit: U/L |
| **Aspartate aminotransferase (AST)** | Unit: U/L |
| **Gamma glutamyltransferase (GGT)** | Unit: U/L |
| **Platelets** | Unit: $x10^9/L$ |
| Bilirubin | Unit: mg/dL |
| International normalised ratio | Unit: ratio |
| Hepatitis B e antigen | Positive/ negative |
| Hepatitis B DNA | Unit: IU/ml |
| Hepatitis B genotype | Genotype assigned from sequencing |
| Anti-hepatitis C antibody | Positive/ negative |
| Hepatitis C RNA | Positive/ negative |
| Anti-hepatitis D antibody | Positive/ negative |
| Hepatitis D RNA | Positive/ negative |
| Body mass index | Unit: $kg/m^2$ |
| **Reason for testing for hepatitis B** | Suspected liver disease, due to clinical features of liver disease, or abnormal liver function tests, or abnormal liver imaging; or asymptomatic screening for antenatal care, or blood donation, or family contact of HBsAg positive individual, or community screening. |

| | |
|---|---|
| **Current or past hepatitis B treatment** | Comprising tenofovir disoproxil fumarate, tenofovir alafenamide, entecavir, lamivudine, emtricitabine, telbivudine, adefovir, interferon. |
| Family history of HCC or cirrhosis | First- or second-degree relative with cirrhosis or HCC. |
| Alcohol abuse | Centre-specific definitions were used. |
| Type 2 diabetes | Ever diagnosed, or treated for type 2 diabetes mellitus. |
| Hypertension | Ever diagnosed, or treated for hypertension. |
| Hyperlipidaemia | Ever diagnosed with, or treated for, hyperlipidaemia. |
| Hepatic schistosomiasis | Evidence of schistosomal liver disease by radiology + a positive serum/stool/urine test (according to centre-specific diagnostics) |
| HCC | Liver tumour(s) diagnosed by radiology or histology. |
| Ascites | Past or current evidence of ascites, by clinical examination and/or radiology. |
| Jaundice | Clinically diagnosed with jaundice by a clinician |
| Variceal bleeding | Upper GI bleeding where endoscopy confirms oesophageal varices. |
| Hepatic encephalopathy | Cerebral dysfunction observed and diagnosed as HE by a clinician. |

Appendix 3: Characteristics of study sites

| Country | Site | Principle investigator(s) | Facility | Year national HBV vaccine introduced | Number of eligible patients | Endemic Schisto-somiasis mansoni | Definition of hazardous alcohol | Biochemistry assay | HBV DNA quantification assay |
|---|---|---|---|---|---|---|---|---|---|
| Ethiopia | Addis Ababa | Desalegn & Johannessen | Referral hospital | 2007 | 1038 | No | WHO AUDIT | Humalyzer 3000, Human | HBV Realtime, m2000sp/rt, Abbott & GeneXpert HBV, Cepheid |
| Gambia | Fajara | Njie & Lemoine | Referral hospital | 1990 | 797 | No | >20g/day (none reported) | VITROS 350, Ortho | In-house assay LLQ=50 IU/ml |
| Senegal | Dakar | Mbaye & Vray | Secondary hospitals (4) | 2005 | 169 | No | Not reported | Not reported | Cobas Ampliprep/Taqman v1.0, Roche |
| Nigeria | Jos | Okeke | Referral hospital | 2004 | 190 | Yes | CAGE questionnaire | Cobas, Roche | In-house assay LLQ=20 IU/ml |
| South Africa | Cape Town | Spearman & Sonderup | Referral hospital | 1995 | 155 | No | WHO AUDIT | Coba 6000, Roche | Cobas Amplicor, Roche |
| Malawi | Blantyre | Stockdale | Referral hospital | 2002 | 97 | Yes | WHO AUDIT | AU480, Beckman Coulter | In-house assay LLQ=35 IU/ml[53] |
| Zambia | Lusaka | Sinkala & Vinikoor | Referral hospital | 2005 | 283 | Yes | WHO AUDIT-C | Multiple platforms | In-house; Cobas Ampliprep/Taqman, Roche & GeneXpert HBV, Cepheid |
| Senegal | Dakar | Fall | Referral hospital | 2005 | 97 | No | WHO AUDIT | Cobas 6000, Roche | Cobas Ampliprep/Taqman v1.0, Roche |
| Senegal | Thies | Lemoine | Referral hospital | 2005 | 300 | No | >20g/day (none reported) | VITROS 350, Ortho | HBV Realtime, m2000sp/rt, Abbott |
| South Africa | Stellenbosch | Maponga | Referral hospital | 1990 | 85 | No | Not reported | Architect, Abbott | HBV Realtime, m2000sp/rt, Abbott |
| Senegal | Dakar | Seydi & Wandeler | Referral hospital | 2005 | 303 | No | Not reported | CYNSTART, Cypress Diagnostics, Belgium | COBAS Ampliprep/TaqMan System, Roche |
| Burkina Faso | Ouagadougou | Sombie | Referral hospital | 2005 | 35 | No | Not reported | Architect ci8000, Abbott | HBV Realtime, m2000sp/rt, Abbott |

Appendix 4: Description of bivariate random effects model

To calculate sensitivity and specificity, data were pooled using a single-stage individual patient data (IPD) meta-analysis approach. We used a bivariate Bayesian random-effects meta-analysis model for sensitivity and specificity using patient-level covariates with study-level random effects to account for anticipated variability between sites.[21]

Specifically, let $Y_{i,j}$ be the random variable recording the outcome for participant $j = 1, \dots, n_i$ in study $i = 1, \dots, m$ for a specific biomarker $X$ and a specific threshold $x_t$ that are currently considered. $Y_{i,j} = 0$ if $X_{i,j} < x_t$ and $Y_{i,j} = 1$ if $X_{i,j} \geq x_t$.

Let $\text{state}_{i,j}$ be the true disease state (according to reference test result, for example cirrhosis present or absent).

The Bayesian bivariate model for sensitivity and specificity is defined by:

$$Y_{i,j} \sim \text{Bernoulli}(p_{i,j})$$

$$\text{logit}(p_{i,j}) = \begin{cases} \beta^{(1)} + \gamma_1^{(1)} \cdot \text{alcohol}_{i,j} + \gamma_2^{(1)} \cdot \text{sex}_{\text{female}_{i,j}} + \gamma_3^{(1)} \cdot \text{BMI}_{\text{underweight}_{i,j}} + \gamma_4^{(1)} \cdot \text{BMI}_{\text{overweight}_{i,j}} + \gamma_5^{(1)} \cdot \text{BMI}_{\text{obese}_{i,j}} + \gamma_6^{(1)} \cdot \text{test reason}_{\text{susp. liver disease}_{i,j}} + u_{1,i} \\ \quad \text{if state}_{i,j} = \text{positive} \\ \beta^{(0)} + \gamma_1^{(0)} \cdot \text{alcohol}_{i,j} + \gamma_2^{(0)} \cdot \text{sex}_{\text{female}_{i,j}} + \gamma_3^{(0)} \cdot \text{BMI}_{\text{underweight}_{i,j}} + \gamma_4^{(0)} \cdot \text{BMI}_{\text{overweight}_{i,j}} + \gamma_5^{(0)} \cdot \text{BMI}_{\text{obese}_{i,j}} + \gamma_6^{(0)} \cdot \text{test reason}_{\text{susp. liver disease}_{i,j}} + u_{0,i} \\ \quad \text{if state}_{i,j} = \text{negative} \end{cases}$$

where

$$p_{i,j} = \begin{cases} P(Y_{i,j} = 1 | \text{state}_{i,j} = \text{positive}) = \text{sensitivity} & \text{if state}_{i,j} = \text{positive} \\ 1 - P(Y_{i,j} = 0 | \text{state}_{i,j} = \text{negative}) = 1 - \text{specificity} & \text{if state}_{i,j} = \text{negative} \end{cases}$$

and $u_{1,i}, u_{0,i}$ are study-specific random effects

$$(u_{1,i}, u_{0,i})^T \sim N((0,0)^T, \Omega)$$

with $\Omega$ a 2x2 covariance matrix.

Since we use a Bayesian paradigm to fit the model, we need to specify prior distributions for the model parameters. These are non-informative priors:

$$\beta^{(l)}, \gamma_k^{(l)} \sim N(0, 10^5) \quad l = 0,1; \; k = 1, \dots, 6$$

and

$$\Omega^{-1} \sim \text{Wishart}\left(\begin{pmatrix} 1 & 0 \\ 0 & 1 \end{pmatrix}, 2\right)$$

In the stratified models, the above model is fitted to the data from each stratum based on testing reason (suspected liver disease or other). For those models, the testing reason terms, i.e. the parameters $\gamma_6^{(l)}, \; l = 0,1$, are dropped in the above model specification.

The model further specifies distributions for all variables in the model: $\text{state}_{i,j}$ (cirrhosis or significant fibrosis present or absent), alcohol, $\text{sex}_{\text{female}}$ are assumed to follow Bernoulli distributions and body mass index (BMI) a categorical distribution. This specification allows the Bayesian model to handle missing values in the dataset: at each MCMC iteration, for the unobserved data values, the model samples from the specified distributions with the corresponding distributional parameters learned from the data. As the model is computationally demanding to fit, we used a grid search with 42 (APRI), 41 (GPR), 41 (ALT) and 43 (FIB4) different threshold values evaluated for each biomarker. We aimed for at least 40 different values per biomarker and the slightly different numbers of thresholds per biomarker results from the fact that for some biomarkers several quantiles have the same value.

Appendix 5: Validation of APRI model for cirrhosis using 500 bootstrap samples

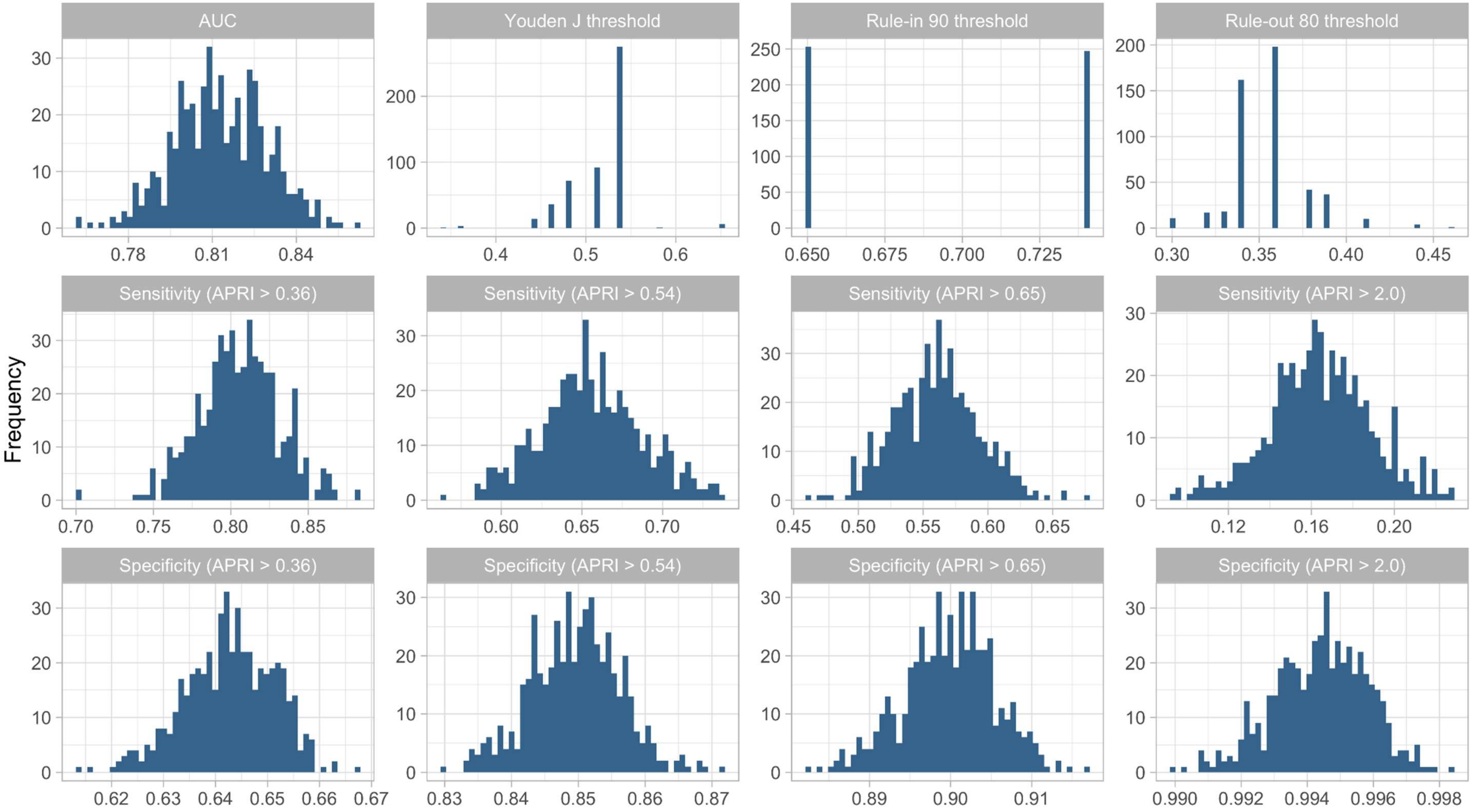

Appendix 6: Ethical approval HEPSANET

Ethiopia
National Research Ethics Review Committee, Ethiopia (Ref.: 3.10/829/07).
Regional Committee for Medical and Health Research Ethics, Norway (Ref.: 2014/1146).

Malawi
National Health Sciences Research Committee of Malawi (Ref.: 16/11/1698 and 15/5/1599).
University of Liverpool, UK (Ref.: 1954).

Zambia
University of Zambia Biomedical Research Ethics Committee (Ref.: 013-09-15).

South Africa
University of Cape Town Human Research Ethics Committee, South Africa (Ref.: 667/2020).
Stellenbosch University Human Research Ethics Committee, South Africa (Ref.: N17/01/013 and S13/04/072).
University of Oxford Tropical Research Ethics Committee, UK (Ref.: OXTREC 01–18).

Nigeria
Jos University Research Ethics Committee, Nigeria (Ref.: JUTH/DCS/ADM/127/XIX/5962).

The Gambia
The Government of The Gambia and Medical Research Council (MRC) Gambia Joint Ethics Committee (Ref.: SCC1266).

Senegal
Senegalese National Health Research Ethics Committee (Ref.: SEN11/34 and SEN19/11).

Burkina Faso
Yalgado Ouédraogo University Hospital Center Institutional Review Board, Burkina Faso (Ref.: 0782).

Appendix 7: Flowchart of searches for eligible studies

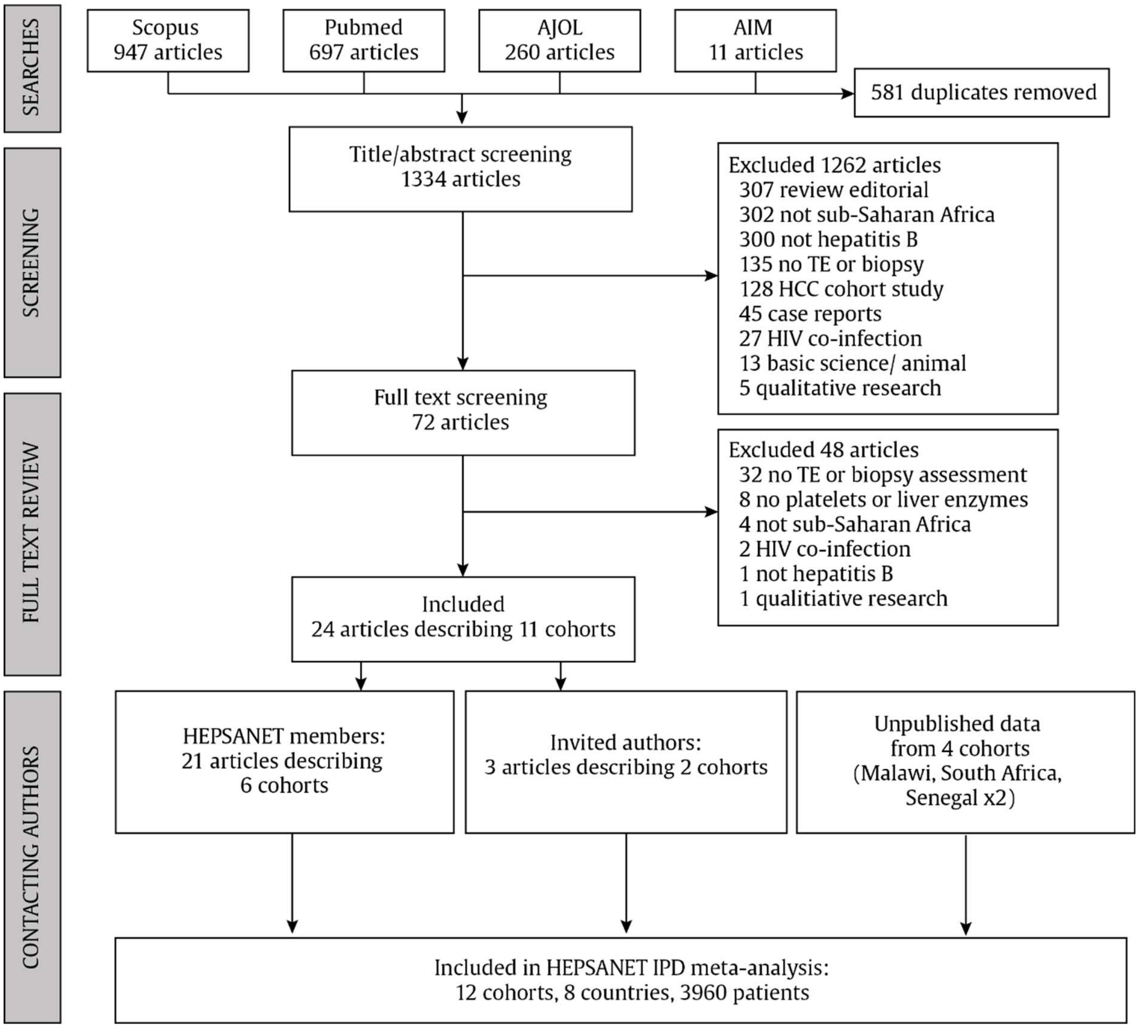

Abbreviations: AJOL, African Journals Online (https://www.ajol.info); AIM, African Index Medicus (https://www.globalindexmedicus.net/biblioteca/aim/); TE, transient elastography; HCC, hepatocellular carcinoma; IPD, individual patient data

Appendix 8: Risk of bias assessment using the QUADAS-2 criteria

| Country | Ethiopia | The Gambia | Senegal 1 | Senegal 2 | Senegal 3 | Senegal 4 | South Africa | South Africa | Nigera | Malawi | Zambia | Burkina Faso |
|---|---|---|---|---|---|---|---|---|---|---|---|---|
| Location | Addis Ababa | Banjul | Theiès | Dakar | Dakar | Dakar | Cape Town | Stellenbosch | Jos | Blantyre | Lusaka | Ouagadougou |
| Setting | Hospital | Community | Hospital | Hospital | Hospital | Hospital | Hospital | Hospital | Hospital | Hospital/ community | Hospital/ community | Hospital |
| **1. PATIENT SELECTION** | | | | | | | | | | | | |
| Was a consecutive or random sample enrolled | Yes | Yes | Yes | Yes | Yes | Yes | No | No[c] | Yes | Yes | Yes | Yes |
| Was a case-control design avoided? | Yes | Yes | Yes | Yes | Yes | Yes | Yes | Yes | Yes | Yes | Yes | Yes |
| Did the study avoid inappropriate exclusions? | Yes | Yes | Yes | Yes | | Yes | Yes | Yes | Yes | Yes | Yes | Yes |
| Could the selection of patients have introduced bias? | No | No | No | Yes[a] | Yes[b] | No | Yes | Yes[c] | Yes[d] | Yes[e] | No | Yes[f] |
| Is there concern that the included patients do not match the review question? | No | No | No | Yes[a] | Yes[b] | No | No | Yes[c] | No | No | No | No |
| **2. INDEX TESTS** | | | | | | | | | | | | |
| Were the index tests interpreted without knowledge of the reference standard? | Yes | Yes | Yes | Yes | Yes | Yes | Yes | Yes | Yes | Yes | Yes | Yes |
| Could the conduct or interpretation of the index test have introduced bias? | No | No | No | No | No | No | No | No | No | No | No | No |
| Is there concern the the index test , its conduct or interpretation differ from the review question? | No | No | No | No | No | No | No | No | No | No | No | No |
| **3. REFERENCE TESTS** | | | | | | | | | | | | |
| Is the reference standard likely to correctly classify the target condiciton? | Yes | Yes | Yes | Yes | Yes | Yes | Yes | Yes | Yes | Yes | Yes | Yes |

| | | | | | | | | | | | | |
|---|---|---|---|---|---|---|---|---|---|---|---|---|
| Were the reference standard results intepreted without knowledge of the results of the index test? | Unclear | Unclear | Unclear | Unclear | Unclear | Yes | No | No | Unclear | Yes | Yes | Unclear |
| Could the reference standard, its conduct, or its interpretation have introduced bias? | No | No | No | No | No | No | No | No | No | No | No | No |
| **4. FLOW AND TIMING** | | | | | | | | | | | | |
| Was there an appropriate interval between index tests and reference standard? | Yes | Yes | Yes | Yes | Yes | Yes | Yes | Yes | Yes | Yes | Yes | Yes |
| Did all patients receive a reference standard? | Yes | Yes | Yes | Yes | Yes | Yes | Yes | No[c] | Yes | Yes | No | No |
| Did patients receive the same reference standard? | Yes | Yes | Yes | Yes | Yes | Yes | No | No | Yes | Yes | Yes | No |
| Were all patients included in the analysis? | Yes | Yes | Yes | Yes | Yes | Yes | No | No | Yes | Yes | No | No |
| Could the patient flow have introduced bias? | No | No | No | No | No | No | Yes | Yes[c] | No | Yes[g] | Yes[g] | No |

[a] Inclusion criteria were inactive HBV carriers with HBV DNA <2000 IU/ml, normal ALT, HBeAg negative.

[b] Inclusion criteria were HBsAg positive for 6 months, treatment naïve, symptom free with HBV DNA >3.2 $log_{10}$ IU/ml.

[c] Subset of patients underwent TE examination at clinicians' discretion - standardised criteria not provided.

[d] Excluded patients with significant alcohol consumption or body mass index >28 $kg/m^2$.

[e] Hospital study recruited patients with suspected cirrhosis based on clinical symptoms or signs suggestive of chronic liver disease.

[f] Only patients undergoing a liver biopsy were included, although this was standard of care at the time for all HBV patients.

[g] Loss to follow up occurred from community diagnosis to treatment eligibility assessment with 94/150 (63%) of HBsAg positive patients being evaluated.

[h] Loss to follow up occurred from referral of patients from the community study to clinical staging at the hospital site with 148/182 (80%) of HBsAg patients having treatment eligibility assessment, of whom 49/148 (33%) had transient elastography.

Appendix 9: Association of cirrhosis prevalence with age, sex, and reason for hepatitis B testing[a]

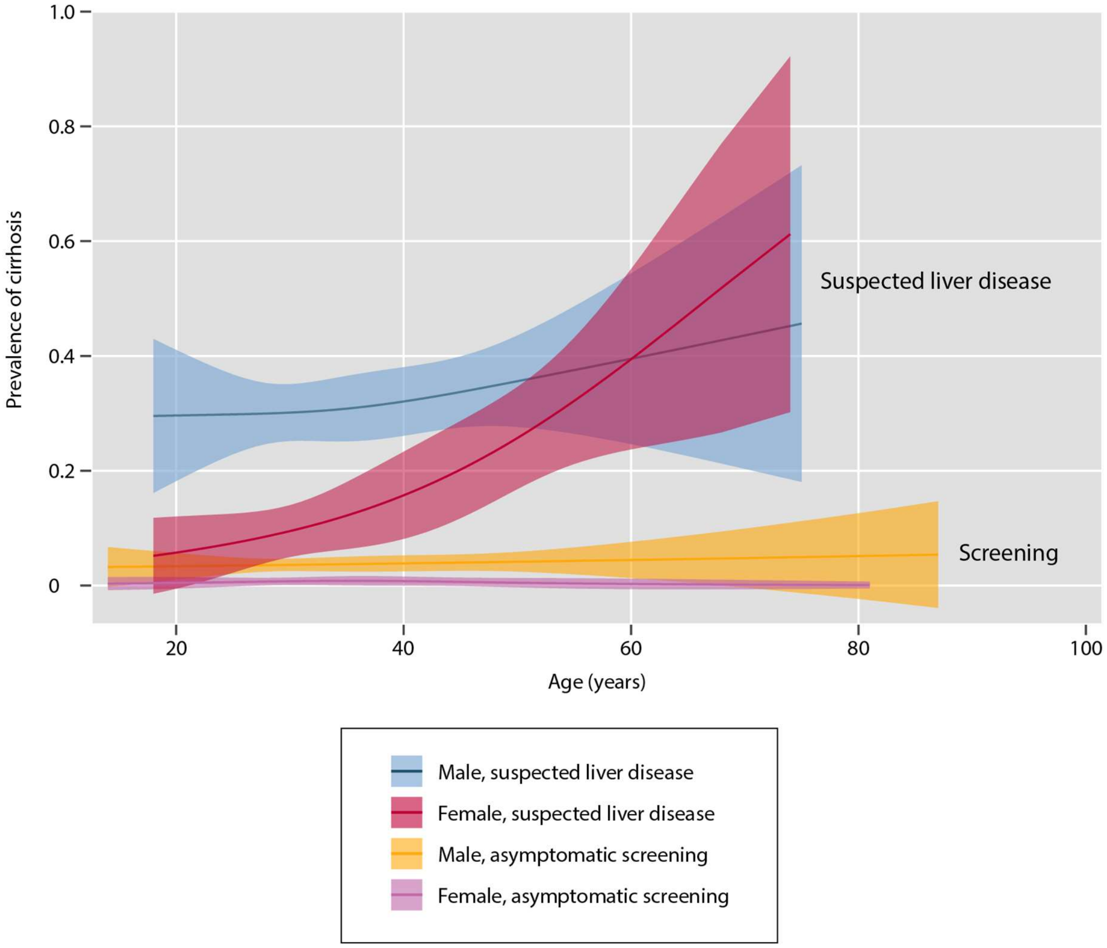

[a] Graphs show restricted cubic splines with three knots with respect to age. Shaded areas surrounding central estimates represent 95% confidence intervals.

Appendix 10: Associations with cirrhosis (model 1) and significant fibrosis (model 2) among HEPSANET participants: mixed effects logistic regression model[a]

**Model 1: Cirrhosis (>12.2kPa)**

| **Variable** | **Univariable association** | | | **Multivariable model** | | |
|---|---|---|---|---|---|---|
| | **Odds ratio** | **(95% CI)** | **P value** | **Odds ratio** | **(95% CI)** | **P value** |
| Age (per year) | 1.03 | (1.02 – 1.04) | <0.001 | 1.03 | (1.01 – 1.04) | <0.001 |
| Sex (male vs female) | 3.55 | (2.52 – 5.00) | <0.001 | 3.31 | (2.19 – 5.00) | <0.001 |
| BMI<br>Underweight<br>Normal<br>Overweight<br>Obese | <br>0.94<br>Reference<br>0.46<br>0.33 | <br>(0.62 – 1.43)<br><br>(0.30 – 0.69)<br>(0.14 – 0.77) | <0.001 | <br>0.90<br>Reference<br>0.58<br>0.45 | <br>(0.56 – 1.45)<br><br>(0.37 – 0.93)<br>(0.17 – 1.21) | 0.06 |
| Suspected liver disease (reference asymptomatic screening) | 45.6 | (25.8 – 80.7) | <0.001 | 55.5 | (28.1 -109.5) | <0.001 |

**Model 2: Significant fibrosis (>7.9kPa)**

| **Variable** | **Univariable association** | | | **Multivariable model** | | |
|---|---|---|---|---|---|---|
| | **Odds ratio** | **(95% CI)** | **P value** | **Odds ratio** | **(95% CI)** | **P value** |
| Age (per year) | 1.01 | (1.00 – 1.02) | 0.022 | 1.01 | (1.00 – 1.02) | 0.07 |
| Sex (male vs female) | 3.38 | (2.70 – 4.23) | <0.001 | 3.42 | (2.63 – 4.45) | <0.001 |
| BMI<br>Underweight<br>Normal<br>Overweight<br>Obese | <br>0.99<br>Reference<br>0.49<br>0.46 | <br>(0.73 – 1.35)<br><br>(0.37 – 0.65)<br>(0.28 – 0.76) | <0.001 | <br>0.92<br>Reference<br>0.60<br>0.65 | <br>(0.66 – 1.30)<br><br>(0.44 – 0.81)<br>(0.37 – 1.12) | 0.005 |
| Suspected liver disease (reference asymptomatic screening) | 8.3 | (6.4 – 10.9) | <0.001 | 9.91 | (7.16 – 13.7) | <0.0001 |

[a]Models include random effects for study site

Appendix 11: Diagnostic performance characteristics at each site, stratified by reason for testing using APRI with rule-out threshold of 0.65 for the diagnosis of cirrhosis

| Site | Asymptomatic screening populations | | | | | Liver disease populations | | | | |
|---|---|---|---|---|---|---|---|---|---|---|
| | Pr (%) | PPV (%) | NPV (%) | Sensitivity (%) | Specificity (%) | Pr (%) | PPV (%) | NPV (%) | Sensitivity (%) | Specificity (%) |
| Ethiopia | 0.6 | 0.0 (0-97.5) | 99.4 (98.6-99.8) | 0.0 (0 - 60.2) | 99.9 (99.2-100) | 36.5 | 61.0 (49.6-71.6) | 72.0 (65.8 - 77.6) | 42.7 (33.6- 52.2) | 84.3 (78.6 - 89.0) |
| Cape Town | 3.7 | 20.0 (2.6-55.6) | 98.0 (92.9-99.8) | 50.0 (6.8 - 93.2) | 92.4 (85.5-96.7) | 8.9 | 16.7 (0.4 - 64.1) | 92.3 (79.1 - 98.4) | 25.0 (0.6 - 80.6) | 87.8 (73.8 - 95.9) |
| Senegal 1 | 0 | - | 100.0 | - | 96.2 (89.3-99.2) | 0.0 | - | 100 | | 100.0 (39.8 - 100) |
| Malawi | 2.7 | 25.0 (0.6-80.6) | 98.5 (92.0-100) | 50.0 (1.3 - 98.7) | 95.7 (87.8 - 99.1) | 91.7 | 95.2 (83.3 - 98.8) | 50.0 (8.6 -91.4) | 95.2 (76.2 - 99.9) | 50.0 (1.3 - 98.7) |
| Nigeria | | | | | | 12.1 | 48.8 (33.3 - 64.5) | 98.6 (95.2 - 99.8) | 91.3 (72.0 - 98.9) | 86.8 (80.7 - 91.6) |
| Stellenbosch | 11.6 | 50.0 (11.8-88.2) | 96.7 (82.8-99.9) | 75.0 (19.4 - 99.4) | 90.6 (75.0 - 98.0) | | | | | |
| Zambia | 2.1 | 0.0 (0-14.8) | 96.2 (87.0-99.5) | 0.0 (0- 84.2) | 68.9 (57.1 - 79.2) | 54.6 | 57.1 (18.4 - 90.1) | 66.7 (9.4 - 99.2) | 80.0 (28.4 - 99.5) | 40.0 (5.3 - 85.3) |
| Gambia | 1.3 | 5.5 (2.2 - 10.9) | 99.5 (98.7-99.9) | 70.0 (34.8 -93.3) | 84.3 (81.6 -86.8) | | | | | |
| Senegal 2 | 9.5 | 32.3 (16.7 - 51.4) | 96.7 (91.7-99.1) | 71.4 (41.9 - 91.6) | 84.7 (77.5 - 90.3) | | | | | |
| Burkina Faso | | | | | | | | | | |
| Senegal 3 | 6.6 | 37.5 (18.8 - 59.4) | 96.4 (93.3-98.3) | 50.0 (26.0 - 74.0) | 94.2 (90.6 - 96.7) | | | | | |
| Senegal 4 | 1.3 | 20.0 (0.5 - 71.6) | 99.1 (96.7-99.9) | 33.3 (0.8 - 90.6) | 98.2 (95.4- 99.5) | 6.9 | 33.3 (0.8 - 90.6) | 93.9 (85.2 - 98.3) | 20.0 (0.5 - 71.6) | 96.9 (89.2 - 99.6) |

**Abbreviations** APRI, aspartate aminotransferase-to-platelet ratio index; Pr, Prevalence of cirrhosis; PPV, positive predictive value; NPV, negative predictive value

Appendix 12: Association between participant characteristics and biomarker sensitivity and specificity for the diagnosis of cirrhosis (>12.2kPa) with APRI and GPR set at rule-in thresholds: Bayesian bivariate random effects model[a]

| **Biomarker, threshold** | **Sensitivity** | **Specificity** |
|---|---|---|
| **Participant characteristics** | **Odds ratio, posterior mean (95% HDI credible interval)** | **Odds ratio (posterior mean) 95% HDI credible interval** |
| **APRI 0.65** | | |
| Hazardous alcohol consumption | 1.19 (0.11 – 2.79) | 0.53 (0.24, 0.86) |
| Underweight | 1.41 (0.43 – 2.67) | 0.92 (0.57 – 1.33) |
| Overweight | 1.27 (0.31 – 2.63) | 1.44 (0.43 – 2.67) |
| Obese | 1.00 (0.00 – 3.10) | 2.15 (0.85 – 3.92) |
| Suspected liver disease | 4.96 (0.67 – 1.67) | 0.13 (0.06 – 0.21) |
| Female sex | 1.66 (0.53 – 3.17) | 2.26 (1.63 – 2.96) |
| Random effects variance (logit) | 0.88 (0.09 – 2.19) | 1.27 (0.35, 2.64) |
| Reference sensitivity/specificity[a] | 0.50 (0.32 – 0.68) | 0.93 (0.91 – 0.95) |
| **GPR 0.47** | | |
| Hazardous alcohol consumption | 3.00 (0.16 – 8.38) | 0.22 (0.07 – 0.41) |
| Underweight | 1.27 (0.24 – 2.66) | 1.08 (0.51 – 1.78) |
| Overweight | 1.31 (0.17 – 3.17) | 0.57 (0.35 – 0.81) |
| Obese | 0.40 (0.00 – 1.48) | 0.72 (0.30 – 1.24) |
| Suspected liver disease | 3.86 (0.39 – 9.85) | 0.32 (0.15 – 0.50) |
| Female sex | 0.93 (0.14 – 1.99) | 2.64 (1.68 – 3.68) |
| Random effects variance (logit) | 1.27 (0.10 – 3.38) | 0.94 (0.20 – 2.10) |
| Reference sensitivity/specificity[a] | 0.60 (0.39 – 0.79) | 0.94 (0.91 – 0.96) |

Abbreviations: HDI, highest density interval; APRI, Aspartate aminotransferase-to-platelet ratio index; GPR, gamma-glutamyl transferase-to-platelet ratio.

[a] Reference category is a male with normal body mass index (18.5-24.9 kg/m$^2$), without hazardous alcohol consumption, with HBsAg testing conducted for asymptomatic screening. We derived odds ratios (for sensitivity and specificity respectively) for the fixed factors included in the bivariate mixed effects logistic regression model. An odds ratio >1 indicates that the corresponding covariate, on average, increases the sensitivity (or specificity), and an odds ratio <1 indicates that the covariate decreases on average the sensitivity (or specificity).

Appendix 13: Sensitivity analyses of rule-in and rule-out thresholds for APRI assessing the effect of exclusion of patients with ascites, use of sex-specific and centre-specific upper limits of normal, and use of an alternative liver stiffness threshold of 9.5 kPa to define cirrhosis

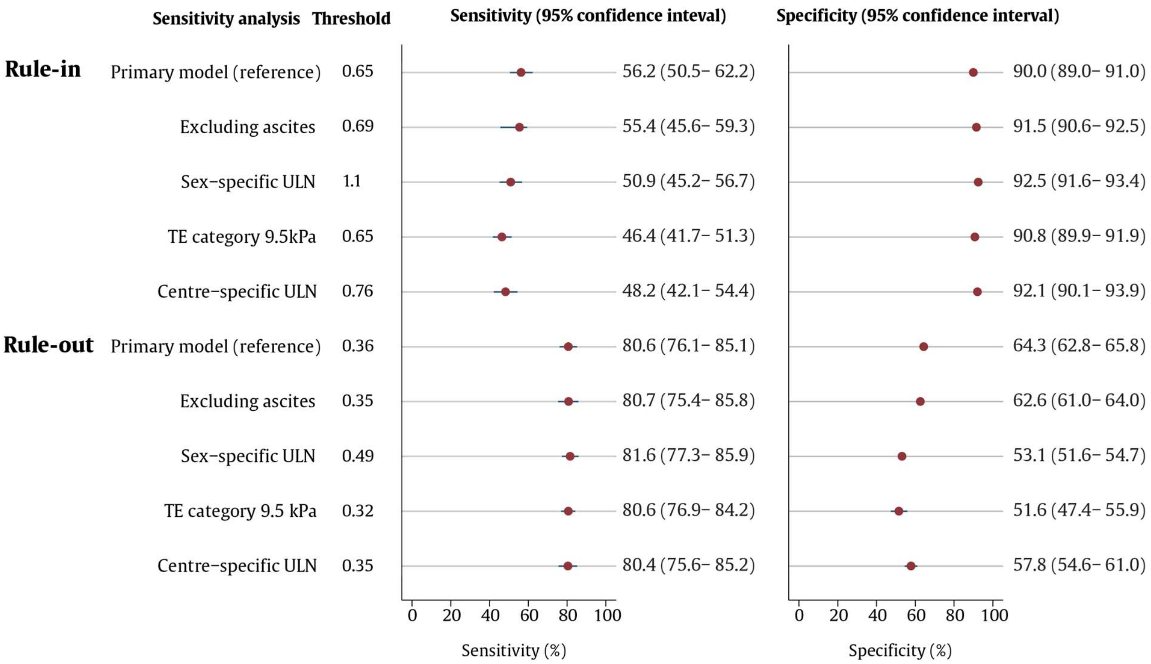

Appendix 14: Association between liver stiffness measurement and test sensitivity for APRI: Liver stiffness distribution stratified by APRI classification (A & B) and sensitivity of APRI relative to liver stiffness (C & D) among patients with cirrhosis

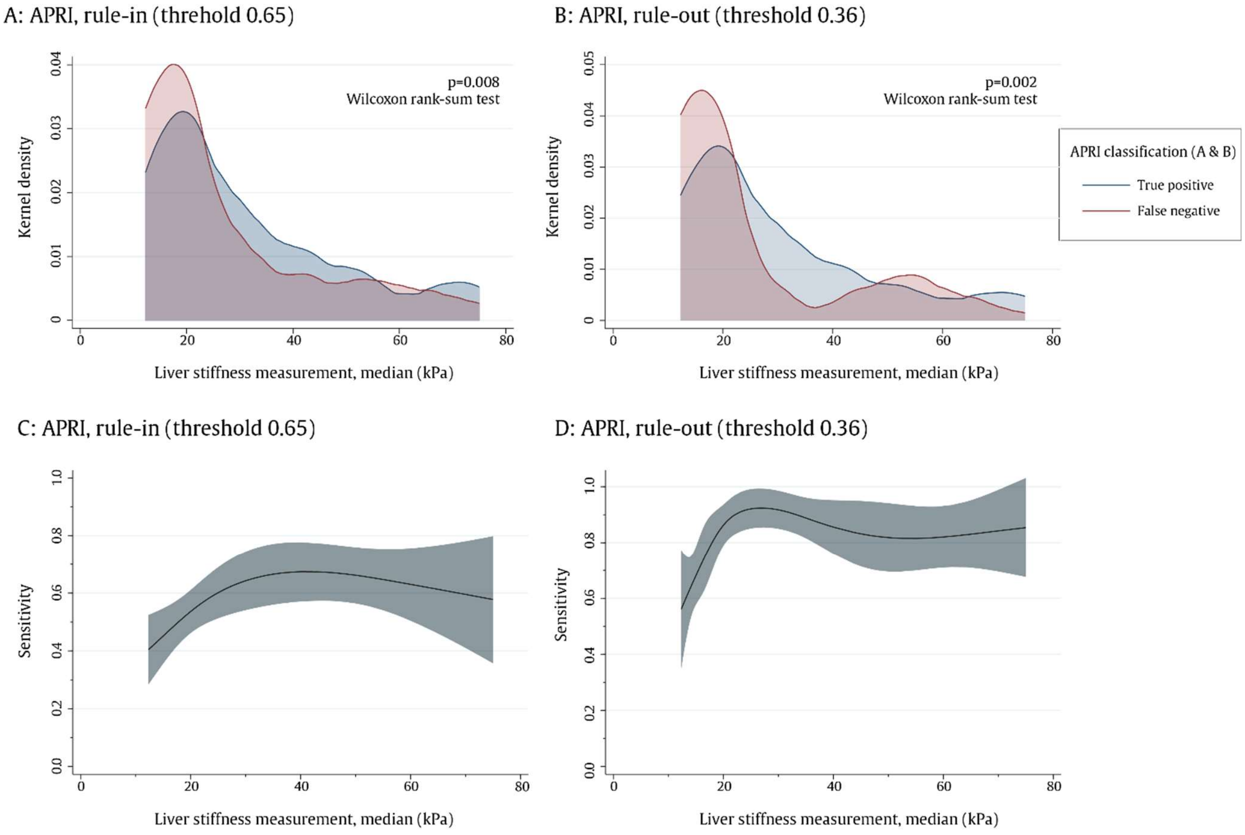

[a] Kernel density plots (A&B) show distribution of median liver stiffness measurements among patients with cirrhosis, stratified by the result of APRI classification at the rule-in (A) and rule-out (B) thresholds. The association between the sensitivity of APRI at rule-in (C) and rule-out (D) thresholds with liver stiffness measurement is shown using a restricted cubic spline with 5 knots, with shaded areas indicating 95% confidence intervals.
